# Factors associated with age at tau pathology onset and time from tau onset to dementia

**DOI:** 10.1101/2025.03.11.25323773

**Authors:** Margo B. Heston, Jordan P. Teague, Karly A. Cody, Yuetiva Deming, Elena Ruiz de Chavez, Jacob Morse, Nathaniel A. Chin, Corinne D. Engelman, Richard J. Chappell, Rebecca E. Langhough, Carey E. Gleason, Lindsay R. Clark, Megan L. Zuelsdorff, Tobey J. Betthauser, Alzheimer’s Disease Neuroimaging Initiative

## Abstract

**INTRODUCTION:** Elevated tau is temporally proximal to dementia onset but less is known about factors influencing T+ onset age and time to dementia following T+ in Alzheimer’s disease. We used sampled iterative localized approximation (SILA) estimated T+ onset age (ETOA) to investigate factors associated with T+ age and time from T+ to dementia onset in ADNI.

**METHODS:** Using SILA-estimated A+ and T+ onset ages derived from ^18^F-Flortaucipir, ^18^F-Florbetapir, and ^18^F-Florbetaben PET and Cox proportional hazards and accelerated failure time models, we analyzed *APOE*, sex, amyloid burden, age, educational attainment, and literacy associations with ETOA and time from T+ to dementia.

**RESULTS:** Higher amyloid, *APOE*-ε4, lower education, and lower literacy associated with younger ETOA. Older ETOA and higher amyloid associated with shorter time from T+ to dementia.

**DISCUSSION:** This work highlights the prognostic value of ETOA and the need to better characterize factors contributing to ETOA and dementia onset in AD.

## 2. Background

Advances in Alzheimer’s disease (AD) biomarkers and therapies have bolstered the need for accurate biomarker-informed dementia risk models. Of crucial importance to slowing or halting disease progression is identifying optimal timing windows for therapeutic intervention. Current longitudinal biomarker studies often describe the “average” AD progression, which follows a stereotypic sequence beginning with amyloid β (Aβ) accumulation, followed by tau aggregation, neurodegeneration, and subsequent cognitive decline and dementia[1]. Recent studies applying new temporal modeling methods[2–6] that produce person-level estimates show there is substantial interpersonal heterogeneity in when people become amyloid positive (A+) and in the time from A+ to dementia onset, even within established risk strata such as *APOE* genotype[5,6]. Anchoring dementia risk to individualized A+ onset age estimates has also shown that *APOE*-ε4 carriage, female sex, and older age of A+ onset accelerate the time from A+ onset to dementia[2,5,6]. Compared to amyloid, tau aggregation is believed to be more proximal to neurodegenerative changes and clinical symptoms [1,7–9], but less is known about factors that explain when tau positivity (T+) occurs and the time from T+ to dementia. A better understanding of factors that explain when tau biomarkers become abnormal and accelerate the time from T+ to dementia is therefore needed to identify maximally effective disease intervention windows and to inform patient prognosis.

Studies have suggested several factors are associated with tau burden and incidence of T+ and dementia and may therefore influence the onset timing of these AD events. For example, prior studies indicate amyloid and *APOE* may affect resistance to tau pathology differently for females and males, and that educational experiences may additionally impact cognitive resilience in the presence of tau pathology (see review [10]). Females have also shown higher tau levels than males, and among A+ individuals females have shown higher regional tau positron emission tomography (PET) burden for a given level of amyloid burden[11], faster cognitive decline [12,13], and stronger associations of *APOE*-ε4 carriage with higher levels[14] and faster accumulation[15] of phosphorylated tau in cerebrospinal fluid (CSF). Beyond biological factors, social determinants of health (SDoH) have been associated with neurodegeneration, brain health outcomes, and cognition [16–20], but SDoH are less studied in the context of amyloid and tau biomarkers. Cognitive resilience studies have reported mixed effects of education on tau deposition and cognitive decline. For example, several studies have indicated lower educational attainment is associated with greater dementia risk[18], but other studies suggest that education is not associated with tau PET binding[21], despite tau deposition being an established risk factor for cognitive decline. For both biological and SDoH factors, it remains unclear if these associations are a result of developing tau pathology at an earlier age, faster progression from T+ to dementia, or a combination of both.

This study builds on our previous work[6,22] that validated sampled iterative localized approximation (SILA) to model longitudinal amyloid PET accumulation, generate person-level estimated A+ onset age (EAOA), and recast tau burden, preclinical cognitive decline and dementia onset and risk in terms of A+ time (i.e., estimated years A+, aka chronicity or amyloid time). This “biomarker clock” framework also enables intuitive communication about risk in terms of years from biomarker positivity rather than in measured units such as standard uptake value ratio (SUVR), which are less interpretable for non-specialists. In this study, we applied SILA to ^18^F-Flortaucipir (FTP) tau PET data from the Alzheimer’s Disease Neuroimaging Initiative (ADNI) to derive individualized estimated T+ onset age (ETOA). We then investigated whether biological and selected SDoH associate with ETOA and the time from ETOA to dementia. Our aims were to 1) characterize the combined influence of amyloid burden, *APOE*, sex, literacy, and educational attainment on ETOA, 2) examine potential predictors of the time from ETOA to dementia in T+ participants, and 3) compare the heterogeneity in time from EAOA to dementia with the heterogeneity in time from ETOA to dementia.

## 3. Methods

### 3.1. Study population and inclusion criteria

Data used in the preparation of this article were obtained from the Alzheimer’s Disease Neuroimaging Initiative (ADNI) database (adni.loni.usc.edu) downloaded from LONI on 30 November 2023. The ADNI was launched in 2003 as a public-private partnership, led by Principal Investigator Michael W. Weiner, MD. The primary goal of ADNI has been to test whether serial magnetic resonance imaging (MRI), positron emission tomography (PET), other biological markers, and clinical and neuropsychological assessment can be combined to measure the progression of mild cognitive impairment (MCI) and early Alzheimer’s disease (AD). ADNI enrollment inclusion/exclusion criteria have been previously published[23–25]. Participants from the ADNI cohort were included in this study if they had available quality-checked preprocessed amyloid and tau PET standard uptake value ratio (SUVR) data and had undergone *APOE* genotyping. Written consent was obtained before enrollment in accordance with the Declaration of Helsinki standards and under Institutional Review Board review for each source study, including the University of Wisconsin Institutional Review Board.

### 3.2. Demographic characteristics and APOE genotyping

Demographic and APOE data were obtained from demographics and APOE results tables. Participant sex of female or male, and race and ethnicity were obtained through self-report. *APOE* genotype, obtained from blood sample analysis[26], was used to index effects of ε2 and ε4 allelic dose. Two *APOE* ε2 homozygotes and 18 *APOE* ε2ε4 individuals were included in depictions of results but were excluded from statistical analysis, due to low participant number.

### 3.3. Cognitive diagnosis and clinical outcomes

Cognitive diagnoses of cognitively unimpaired, MCI and AD dementia established by the ADNI as previously described[27] were used for participant summaries but were not further analyzed. An AD dementia diagnosis required meeting the National Institute of Neurological and Communicative Disorders and Stroke–Alzheimer’s Disease and Related Disorders Association criteria for probable AD[28]. The Clinical Dementia Rating (CDR)[29] global score and sum of boxes were used to investigate dementia onset and clinical progression. For primary analyses, the age of first CDR global score ≥1 was used to establish dementia onset and dementia onset age. Sensitivity analyses also investigated MCI onset and onset age, using the first age for which two consecutive visits had CDR global score ≥0.5 as a proxy for MCI. Two abnormal CDR visits were required because we observed that ∼7% (14/190) of those with CDR global scores of 0.5 reverted to 0 at the next observation. Clinical trajectories were depicted continuously using CDR Sum of Boxes (CDR-SB) scores, using the first observation of CDR-SB≥4.5 (equivalent to CDR global score≥1) to define dementia onset and dementia onset age[30].

### 3.4. Social determinants of brain health

SDoH variables representative of educational experiences were obtained from the demographics and neuropsychological battery tables. Specifically, we selected education and literacy measures due to strong evidence that these are associated with dementia outcomes[31–33]. Further, these variables were available for most ADNI participants with tau PET data, and are collected across many biomarker studies, allowing for future replication in additional cohorts. Educational attainment was measured using self-reported years of completed education; we stratified attainment levels by high school education or less (≤12 years completed), some college (13-16 years), and postgraduate education (>16 years). Literacy was measured using the American National Adult Reading Test (ANART); it should be noted that the ANART is not an indicator of premorbid intelligence and is instead a proxy for early life educational quality and literacy-building enrichment in adulthood[31,33]. The ANART is a 50-item scale that indexes performance by summing a participants’ errors when reading words with atypical phonemic pronunciation[34,35]; more total reading errors indicates lower exposure to literacy enrichment through early life education and adulthood occupation[36,37].

### 3.5. PET image processing, quantification, and positivity

Imaging biomarker outcomes were obtained from the UC, Berkeley Amyloid PET and Tau PET analysis tables on LONI. PET images were processed and quantified at the University of California, Berkeley using previously published MR-guided pipelines with FreeSurfer parcellations to derive SUVRs in subject space[38,39]. SUVRs from ^18^F-Florbetapir (FBP; ADNI GO, 2, 3) and ^18^F-Florbetaben (FBB; ADNI 3) scans were used to measure brain amyloid burden, and FTP (ADNI 3) were used to measure tau burden.

#### 3.5.1. Amyloid PET processing and quantification

Cortical amyloid burden was quantified using late-frame FBP (SUVR, 50-70 minutes post-injection) and FBB (SUVR, 90-110 minutes post-injection) amyloid PET imaging. Cortical SUVR was averaged across 20 frontal, anterior/posterior cingulate, lateral parietal, and lateral temporal regions and normalized to a composite reference region comprising the whole cerebellum, brainstem, and eroded white matter (Supplementary Figure 1A), as this has been shown to improve longitudinal amyloid PET trajectories in ADNI[40]. Cortical SUVRs were then converted to Centiloids (CL) using previously published linear transforms[41]. Amyloid CL was included in statistical models continuously, as specified in Section 3.3, and was stratified into A-(<19 CL), moderate amyloid (19-75 CL), and high amyloid (>75 CL) groups to depict the effect of amyloid burden on ETOA and the time from T+ onset to dementia.

#### 3.5.2. Tau PET processing and quantification

Tau burden was quantified using late-frame FTP imaging (SUVR, 80-100 minutes post-injection) normalized to inferior cerebellar gray matter. Analyses used a meta-temporal FTP SUVR average across the bilateral entorhinal cortex, amygdala, fusiform gyrus, inferior temporal gyrus, and middle temporal gyrus (Supplementary Figure 1B).

#### 3.5.3. Amyloid and tau positivity thresholds

We applied Gaussian mixture modeling with two classes separately to baseline amyloid PET CL and baseline tau PET SUVR values (Supplementary Figure 2A-B). For each PET biomarker, the positivity threshold was calculated as the mean plus two standard deviations (SD) above the lower mean component (i.e., the presumed biomarker negative group). This yielded an A+ threshold of 19 CL and a meta-temporal T+ threshold of 1.36 SUVR (Supplementary Figure 2C-D).

### 3.6. Amyloid and tau trajectory modeling and estimated A+ and T+ ages

We applied sampled iterative local approximation (SILA; https://github.com/Betthauser-Neuro-Lab/SILA-AD-Biomarker) separately to the CL and FTP SUVR data to model longitudinal biomarker trajectories, derive person-level estimated A+ and T+ onset ages (EAOA and ETOA, respectively), and estimate amyloid burden (i.e., CL) at ETOA and amyloid and tau burden (i.e., CL and FTP SUVR) at CDR observations (Supplementary Figure 2E-F). The details of the SILA method have been explained previously[6]. Briefly, for each biomarker, SILA was trained on all participants with longitudinal observations for that biomarker to define an SUVR (or CL) vs. biomarker time curve with the initial condition that time=0 years at the biomarker positivity threshold. Basis functions were then used to identify the optimal time shift (Δt) for each person that aligns their observed SUVR/CL vs. age data onto the modeled SUVR/CL vs. biomarker time curve; the optimal solution minimizes the within-subject sum of squared SUVR/CL residuals. ETOA and EAOA were then calculated for each person by subtracting their mean estimated biomarker positive times from the mean of their age(s) at scan(s). To verify amyloid trajectories did not differ by PET tracer, we first applied SILA to FBP and FBB CL data separately and compared the CL vs. A+ time curves (Supplementary Figure 2G). After seeing strong concordance between FBP and FBB trajectories, we applied SILA to the combined FBP and FBB CL data for final analyses. In addition to estimated biomarker times and onset ages, SILA was also used to estimate cortical amyloid burden at ETOA by solving the modeled CL vs. biomarker time function for CL at the time corresponding to ETOA (or last T-scan for T-‘s) for each participant. This approach was repeated to estimate cortical amyloid and meta-temporal tau burden at each CDR timepoint.

### 3.7. Statistical analysis

All statistical analyses were performed using R software version 4.3.2 and the survival package (3.5-8). Kaplan-Meier curves were plotted with ggsurvfit (1.0.0), ROIs were visualized using ggseg (1.6.5), and participant statistics were summarized using gtsummary (1.7.2); all other visualizations used ggplot2 (3.5.1), ggh4x (0.2.8), and patchwork (1.2.0). A forward model selection process was used to identify significant explanatory variables and is described for Aims 1 and 2 below. A CONSORT diagram summarizing inclusion criteria and datasets for each aim is provided in Figure 1.

**Figure 1.**
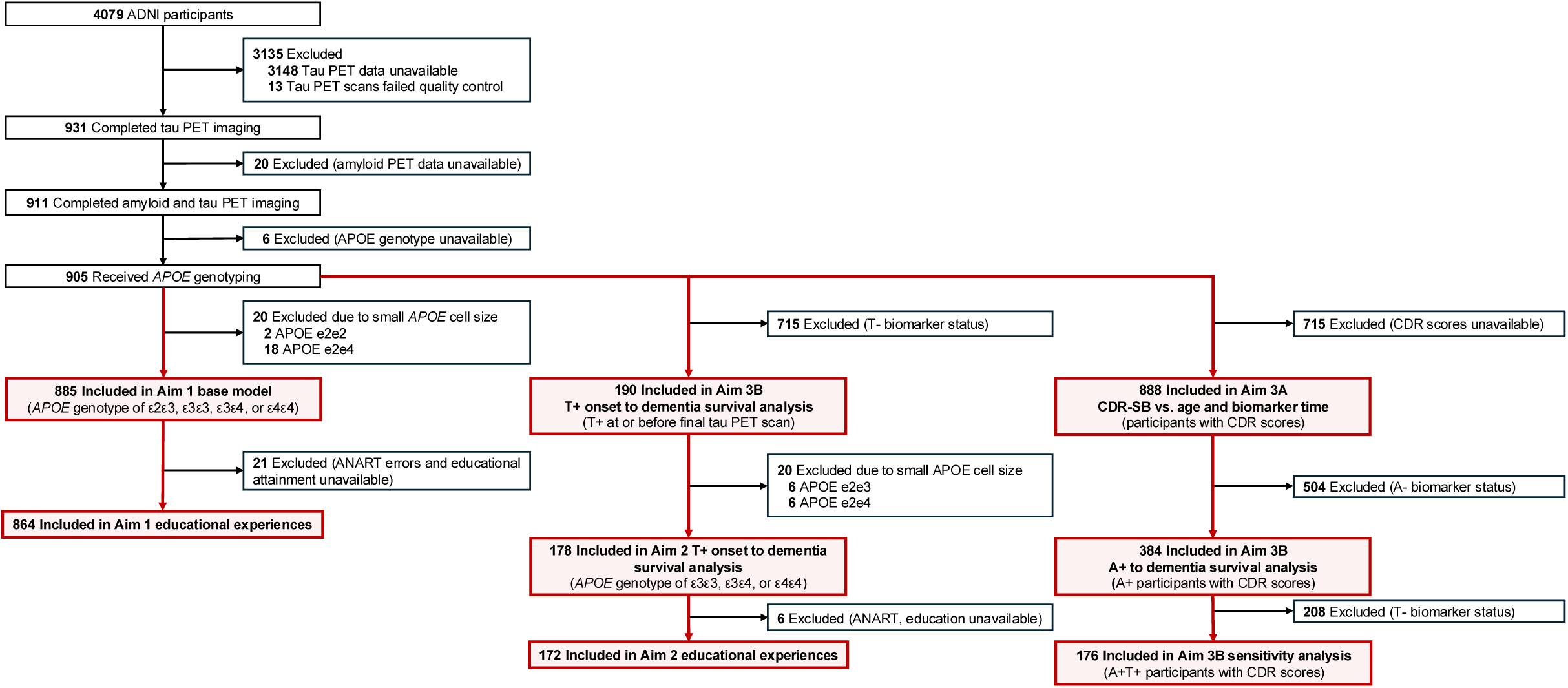
Selection of analysis datasets per study aim. Flow diagram indicating criteria for inclusion in study analyses. Final analysis datasets are highlighted in red and labeled per aim. For Aims 1 and 2, participants with *APOE* genotypes that were insufficiently represented in the dataset were excluded from analysis due to small cell size. Abbreviations: A±, amyloid positive or negative status assessed using ^18^F-Florbetapir or ^18^F-Florbetaben PET neuroimaging; ANART, American National Adult Reading Test; *APOE*, apolipoprote=in E; CDR, Clinical Dementia Rating; T±, tau positive or negative status assessed using ^18^F-Flortaucipir PET neuroimaging.

#### 3.7.1. Aim 1: *APOE*, sex, amyloid, and educational experiences as predictors of ETOA

885 participants comprised the analysis cohort included in Aim 1 (Figure 1). Cox proportional hazards regression was used to evaluate the effects of biological factors and educational experiences on T+ risk. The model used ETOA as the event of interest for T+ individuals with right-censoring at age of last FTP scan for participants who remained tau negative (T-). Biological predictors first included main effects of sex, *APOE* genotype, and amyloid burden estimated at ETOA (or censor), and age at baseline FTP scan as a covariate. Additionally, we tested prespecified exploratory models that included *APOE*-by-sex and amyloid-by-sex interactions, as well as ANART total errors and educational attainment main effects. *APOE* genotype was modeled as a 4-level categorical variable using an ε3ε3 reference contrast (excluding *ε2ε2* and *ε2ε4* genotypes), with *APOE-ε3ε3* males were the reference group when testing the interaction of *APOE*-by-sex. Amyloid was modeled continuously with 0 CL as the reference value with hazard ratios reported depicting 10 CL increases in amyloid burden. Age at baseline FTP was modeled continuously and hazard ratios are reported as risk per 10-year age increments. Starting with main effects, we used forward selection to add model terms in the order listed above and identify those that significantly explained variance in ETOA. For each of four selection rounds we assessed goodness of model fit with the likelihood ratio test, Akaike information criterion (AIC) and Bayesian information criterion (BIC). We excluded terms that did not significantly decrease the model’s log-likelihood or increased the AIC or BIC, and arrived at a base model of ETOA = sex + *APOE* + (amyloid at ETOA) + (age at baseline FTP). After base model specification, we tested *APOE*-by-sex and *APOE*-by-amyloid interactions separately and in the same model. In 864 participants with data on educational experiences, we tested main effects of ANART total errors and educational attainment. Because literacy enrichment starts in early childhood and continues throughout schooling and occupational experience in adulthood, ANART reading error effects were first evaluated alone, and then with the effect of high school, college, or postgraduate education added to the model. For each variable we reported exponentiated hazard ratios with 95% confidence interval [HR (95% CI)] per risk stratum and calculated *P*-values using Wald’s test; significant effects met a two-sided alpha of 0.05. To account for heterogeneous participant follow-up time across the sample, we conducted a sensitivity analysis using age at each tau scan as a time dependent covariate, instead of controlling for age at baseline scan. Stratified Kaplan-Meier curves were generated using survminer[42] to illustrate significant effects identified in the base model and with addition of educational experiences.

#### 3.7.2. Aim 2: Effects of biological factors and educational experiences on time from T+ onset to dementia

Aim 2 analyses were restricted to T+ participants (n=178, Figure 1). We included *APOE* genotypes represented by at least twenty-nine people (ε3ε3 n=59, ε3ε4 n=90, ε4ε4 n=29) and included sixteen A-T+ participants who were later excluded in sensitivity analyses. Time to dementia was estimated as age at first CDR global score ≥1 minus ETOA for those with CDR global score ≥1, and age at last CDR observation minus ETOA for those who remained CDR global score <1 across all observations (i.e., we right censored time for those who did not progress to dementia, n=100). Because some participants had dementia (n=36) up to 0.24 years prior to ETOA and the model requires positive time, we applied a linear shift in the time to event variable, adding 0.24 years. Using an approach similar to Aim 1, we used accelerated failure time (AFT) models to identify factors that predict the time from T+ onset to dementia. Specifically, conducting forward model selection, we sequentially tested the main effects of amyloid burden at ETOA (continuous CL, divided by 10 as in Aim 1), ETOA itself (continuous years, divided by 10 to reflect 10-year increments in ETOA), *APOE* genotype (categorical with ε3ε3 reference group), sex (dichotomous with male reference group), and age at baseline FTP scan (continuous years, divided by 10 as in Aim 1), retaining predictors that significantly reduced the model’s log-likelihood and did not increase AIC or BIC. ETOA was included as a predictor to build on prior work showing that EAOA influences the time from amyloid onset to dementia[6]. We hypothesized that ETOA similarly affects the time from T+ to dementia. Model specification yielded a base model of (time from T+ onset to dementia) = (amyloid at ETOA) + ETOA. After forward selection, we tested *APOE*-by-sex (ε3ε3 males as reference group) and amyloid-by-sex (males with 0 CL as reference group) interactions in prespecified exploratory models. Educational attainment and ANART errors were examined sequentially after examining the interactions. Effects were reported as the difference in time from T+ to dementia across groups and were expressed in years (e.g., a coefficient of −0.10 represents a 10% faster progression from T+ to dementia compared to the contrast group). To provide more interpretable survival time estimates in visualizations, untransformed time to event was used to calculate median survival times and display Kaplan-Meier curves for significant effects.

#### 3.7.3. Aim 3: Comparison of EAOA and ETOA as predictors of time to dementia

Aim 3A: 888 participants with amyloid, tau, and CDR were included in analysis for Aim 3A (Figure 1). We used CDR-SB to examine continuous relationships of cognitive decline with respect to biomarker timelines, and we calculated A+ and T+ time at each CDR timepoint as (age at CDR – EAOA) and (at age CDR – ETOA), respectively. CDR-SB trajectories were then plotted for all participants to visualize heterogeneity along chronological age, A+ time, and T+ time. Dementia onset ranges along each timeline were approximated by calculating the range of participants’ ages and biomarker times at first CDR-SB≥4.5.

Aim 3B: To directly compare the effects of biomarker onset age on time from A+ and T+ onset to dementia, we applied accelerated failure time models as in Aim 2 and estimated risk of dementia at 5-year intervals (5 years through 25 years) after biomarker onset, stratifying risk by decade of EAOA and ETOA for their respective timelines. Models included 384 A+ participants and 190 T+ participants, respectively (Figure 1). Dementia was defined as global CDR≥1. Time to event was the difference between age at dementia onset and biomarker onset age, censored as in Aim 2. Models were also retested for MCI risk (CDR≥0.5 at two consecutive observations) and with MCI or dementia risk stratified by decade of baseline CDR age, as sensitivity analyses.

Analyses and visualizations for Aim 3 were repeated in the A+T+ subset (n=176, Figure 1) to assess whether times from amyloid and tau onset to dementia were sensitive to disease severity (i.e., whether including A+T- or A-T+ participants significantly altered the probability estimates of converting to dementia).

### 3.8. Data availability

ADNI data are available upon request at https://adni.loni.usc.edu/data-samples/access-data/. Code for the SILA algorithm is available at https://github.com/Betthauser-Neuro-Lab/SILA-AD-Biomarker.

## 4. Results

### 4.1. Study sample participant demographics

Participant summary statistics and demographic information are provided in Table 1 and Supplementary Table 1. Across the full sample, participants were 51% (464/905) female with a mean (SD) baseline age of 71.2 (7.0) years. Forty-three percent (379/905) of participants had baseline clinical diagnoses of MCI or AD dementia (n=22 missing diagnoses) and 39% (352/905) were *APOE-ε4* carriers. Forty-four percent (399/905) were A+ at or before their last amyloid PET scan with the A+ group having mean (SD) EAOA of 66.3 (9.9) years. Twenty-two percent (198/905) were T+ by their last tau PET scan with T+ participants having mean (SD) ETOA of 72.0 (9.5) years. Most participants identified as non-Hispanic (93%), White (85%), and were highly educated (46% with postgraduate education). ANART reading errors (Supplementary Figure 3) averaged 11.6 (9.7) across the cohort and varied significantly across baseline diagnosis (CU: 10.0 (9.3), MCI: 13.2 (9.8), AD: 16.0 (10.0)) and educational attainment (high school or less: 20.1 (9.5), some college: 12.7 (9.7), postgraduate: 8.7 (8.4)).

**Table 1.**
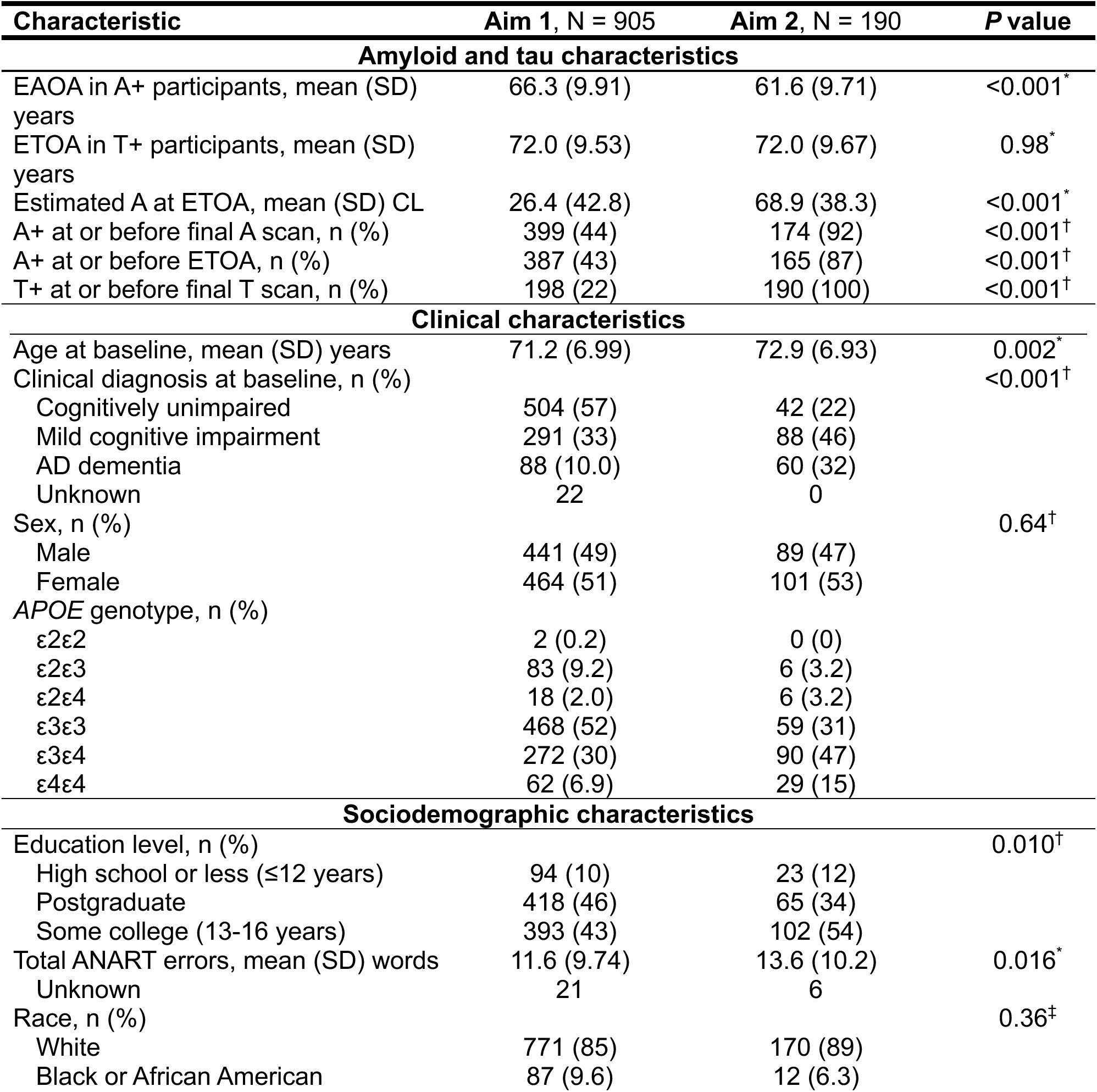
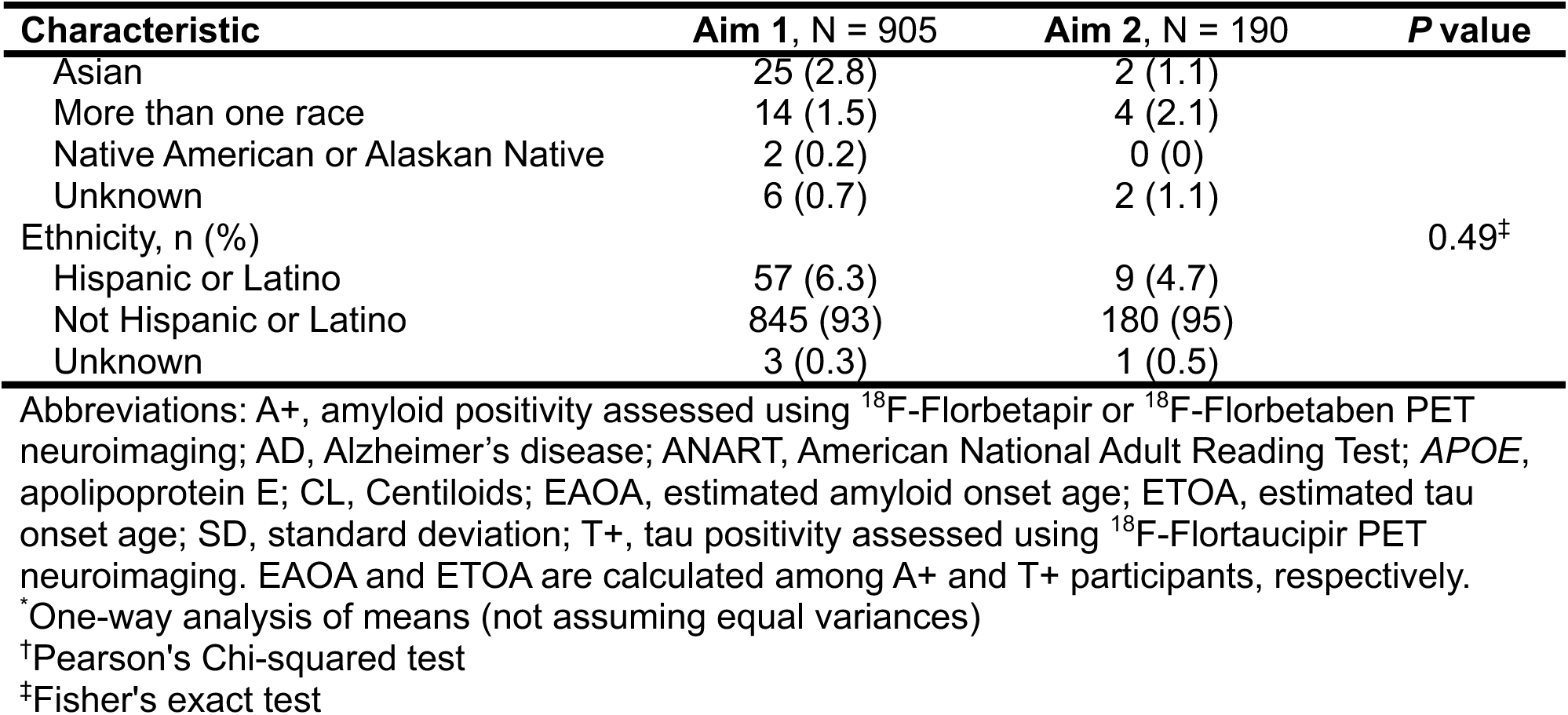
Demographics for participants included in Aim 1 and 2 analyses.

Analysis subsets were demographically representative of the full cohort, but the T+ subset in Aim 2 had significantly higher ANART reading errors (mean (SD)=13.6 (10.2)), and a lower proportion of postgraduate-educated participants (34%). In addition, A+ and T+ subsets were also 1.5-1.7 years older at baseline, had higher rates of cognitive impairment (57% and 78%, respectively), and higher levels of amyloid at ETOA (66.2 (33.7) and 68.9 (38.3) CL, respectively). T+ participants had a younger average EAOA (61.6 (9.7) years) compared to the full cohort.

### 4.2. Aim 1: Factor effects on ETOA

Forward model selection indicated the best fitting Cox proportional-hazards model comprised main effects of sex, *APOE* genotype, and amyloid burden at ETOA, and age at baseline FTP scan (Figure 2A, Supplementary Table 2). Neither *APOE*-by-sex, amyloid-by-sex, nor both interactions together (interaction Wald *p*=0.22, 0.95, 0.36, respectively) significantly improved the model (Supplementary Table 2, Models 1.5, 1.6, 1.7).

**Figure 2.**
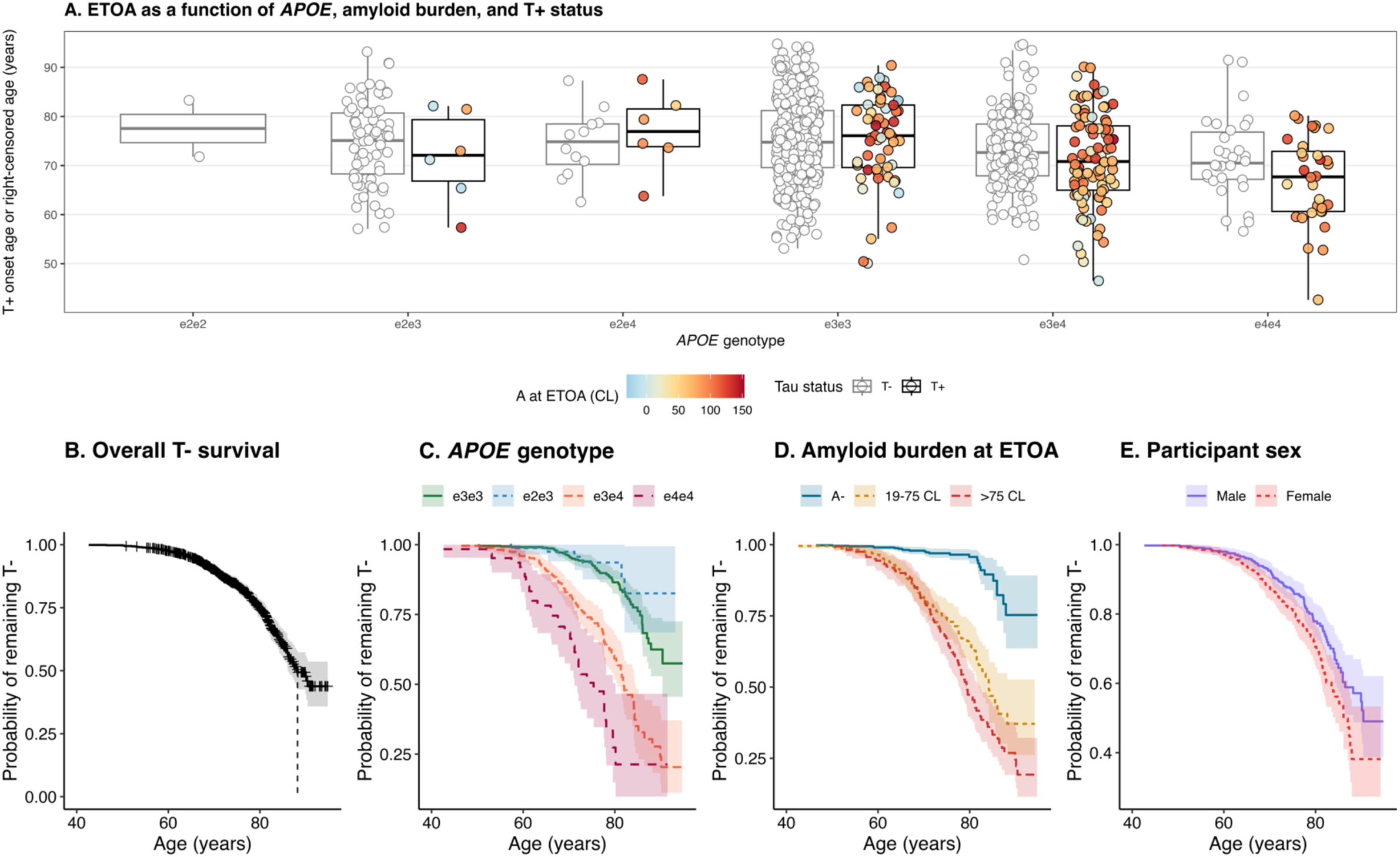
Distribution of ETOA and factors associated with tau onset age. (**A**) Estimated age at T+ in the meta-temporal ROI across *APOE* genotypes with plot color indicating estimated amyloid burden at ETOA for T+ cases. Age at censor is shown with gray circles for T-. (**B-E**) Kaplan-Meier curves depicting the probability of remaining T-as a function of age across levels of main effects terms included in the Cox proportional hazards base model. Abbreviations: A-, amyloid negative status assessed using ^18^F-Florbetapir or ^18^F-Florbetaben PET neuroimaging; *APOE*, apolipoprotein E; CL, Centiloids; ETOA, estimated tau onset age; T±, tau positive or negative status assessed using ^18^F-Flortaucipir PET neuroimaging.

#### 4.2.1. Amyloid, APOE, sex and age

Median (95% CI) T-survival age evaluated with Kaplan-Meier analysis was 88.2 years (lower 95% limit=86.1; upper limit was inestimable; Figure 2B). The *APOE-*ε4 allele showed a dose-dependent effect on T+ risk and ETOA. Relative to the ε3ε3 group, ε3ε4 and ε4ε4 groups had significantly higher T+ risk (HR [95% CI]=1.78 [1.27, 2.51] and 2.71 [1.70, 4.33], respectively; *p*<0.001 for both) with median [lower/upper 95% limit] ETOA of 81.9 [80.6, 84.3] years and 73.9 [71.2, 79.6] years, respectively (Figure 2C). Median T-survival times for ε3 homozygotes could not be estimated because this group did not reach 50% T+. While not significant (*p*=0.69), the ε2ε3 group had lower T+ risk (HR=0.84 [0.36, 1.97]; median T-survival was inestimable). Higher amyloid burden at ETOA conferred greater T+ risk (Figure 2D, HR=1.21 [1.17, 1.25], *p*<.001). T-survival stratified by amyloid groups indicated earlier T+ onset age for those with higher amyloid at T+ onset; median ETOA was 84.2 years (lower 95% limit=81.9) among participants with 19-75 CL and was 79.3 [77.8, 82.3] years among those with >75 CL at ETOA. In T+ participants, median (range) of A+ duration was 6.8 (0.7, 9.8) years and 14.3 (9.9, 23.0) years for those with 19-75 CL and >75 CL at ETOA, respectively. Females had a significantly higher T+ risk than males before but not after controlling for baseline age (Figure 2E, age-adjusted HR [95% CI]=1.11 [0.80, 1.52], *p*=0.097). Older baseline age was significantly associated with lower T+ risk (HR=0.12 [0.09, 0.17] per 10-year increment, *p<*0.001). Stratifying by decade of baseline age indicated participants 75-85 years old at baseline had a greater T+ risk (median ETOA=85.2 years, lower 95% limit=84.3) compared to the younger and older groups, which did not reach 50% T+. Sensitivity analyses controlling for time dependent age at tau scan preserved the direction and significance of effects across biological variables and educational experiences.

#### 4.2.2. Education attainment and literacy exposure

Adding literacy exposure and education attainment variables to the base model significantly improved ETOA model fits, both when included separately and together in the same model. Coefficients here are reported from the model including both variables together; see Supplementary Table 3 for the separate models. Participants with more ANART errors had a significantly greater T+ risk (HR=1.022 [1.009, 1.036], *p*<0.001; Supplementary Table 3) and became T+ earlier (median [lower 95% limit] ETOA in years for 0-20 errors=89.9 [86.5]; 20-30 errors=87.0[81.5]; 30-50 errors=82.8 [77.2]; Figure 3A). ANART effects remained significant after including education attainment (HR=1.019 [1.004, 1.034], *p=*0.011). Education was also significantly associated with ETOA (Figure 3B). Compared to the college-educated reference, participants with postgraduate education had lower risk of having tau pathology (HR=0.69 [0.49, 0.96], *p=*0.027; Supplementary Table 3), whereas those with high school education or less did not differ significantly from the college-educated group (HR=0.83 [0.52, 1.32], *p*=0.43).

**Figure 3.**
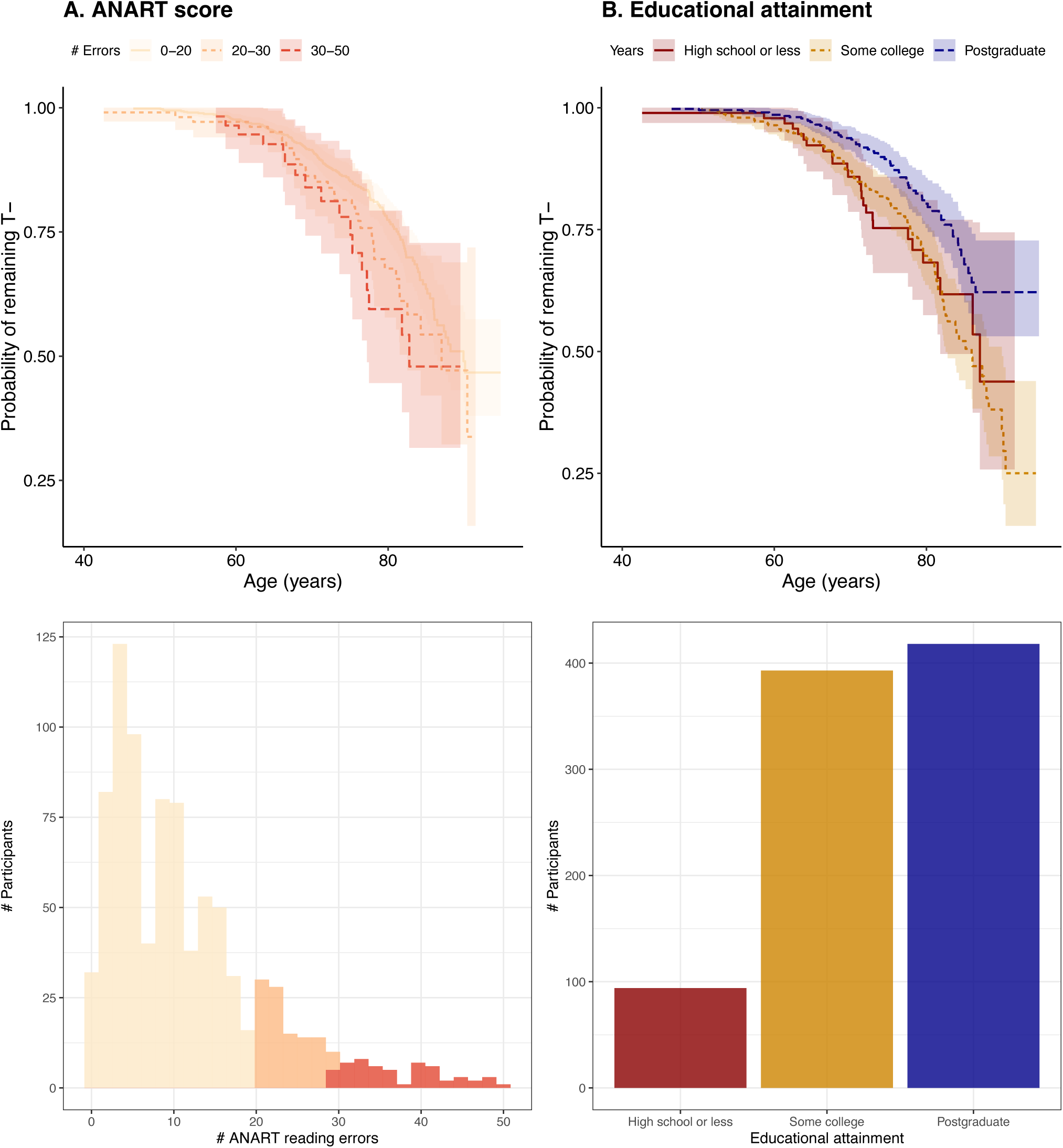
SDoH effects on ETOA. Kaplan-Meier curves depicting the probability of remaining T-as a function of age (top), and participant distributions (bottom) shown across levels of (**A**) ANART reading errors and (**B**) education attainment. SDoH predictors explained significant variance beyond biological effects in the Cox proportional hazards base model. Education attainment levels are defined as: high school or less (≤12 years), college (13-16 years), postgraduate (>16 years). Abbreviations: ANART, American National Adult Reading Test; SDoH, social determinants of health; T-, tau negative status assessed using ^18F^Flortaucipir PET neuroimaging.

### 4.3. Aim 2: Factors associated with time from T+ to dementia

The best fitting AFT model included main effects of ETOA and amyloid burden at ETOA on time from T+ to dementia (Supplemental Table 4). ETOA effects are reported per 10-year increment and amyloid effects are reported per 10 CL increase. Within the T+ subset (n=190), median time from T+ to dementia was 7.48 [6.65, 8.82] years, with only one participant remaining dementia-free 14.1 years after T+ onset who converted to dementia 19.9 years after T+ onset (Figure 4A). Higher amyloid burden at ETOA was associated with shorter time from T+ to dementia onset (*β* [95% CI]=0.046 [−0.079, −0.012], *p=*0.009; Supplementary Table 4). This effect was similar when excluding 16 A-T+ individuals from analysis. Stratifying by amyloid CL showed median times from T+ to dementia were 10.0 [8.89, 14.1] years, 7.48 [lower 95% limit 5.98] years, and 5.76 [3.86, 7.64] years for the A-, moderate amyloid, and high amyloid groups, respectively (Figure 4B). Older ETOA was also associated with shorter time from T+ to dementia (*β* [95% CI]*=*0.29 [−0.42, −0.15], *p<*0.001; Figure 4D). Kaplan-Meier curves (Figure 4C) stratified by ETOA groups depict the ETOA effect on time from T+ to dementia with median times from T+ to dementia=9.13 [7.87, 10.6], 8.58 [lower 95% limit=6.41], 5.98 [5.76, 7.48], 3.07 [lower limit=2.67] years for ETOA <60 years, 60-70 years, 70-80 years, and >80 years, respectively. Neither ANART reading errors (*p*=0.091) nor educational attainment (*p*=0.58) were significantly associated with time from T+ onset to dementia (Supplementary Table 5).

**Figure 4.**
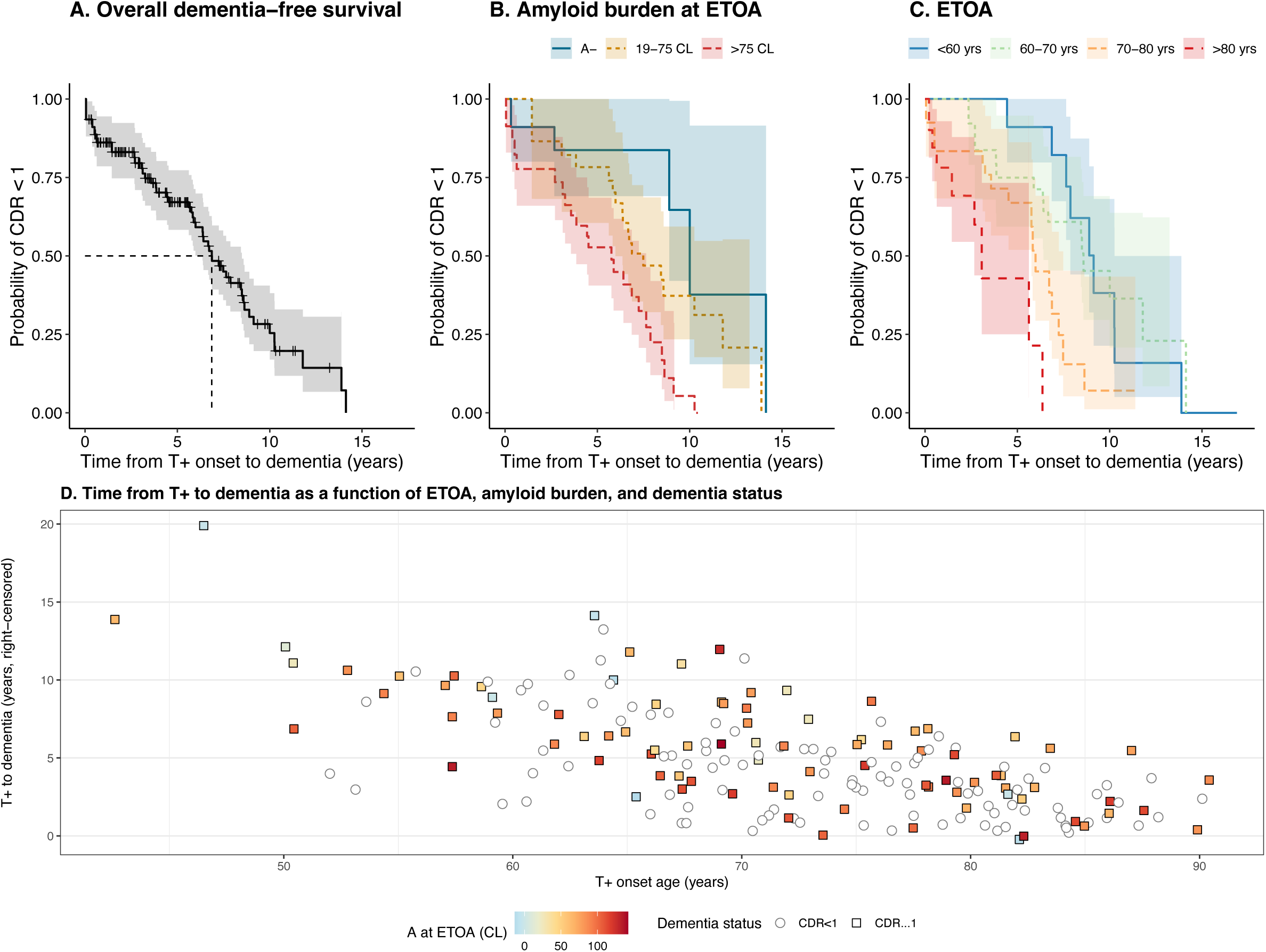
Factors associated with time from T+ to dementia onset. (**A-C**) Kaplan-Meier curves comparing survival probabilities across levels of significant predictors of dementia onset risk. Models were evaluated among T+ participants, excluding twelve total participants with ε2ε2, ε2ε4, and ε2ε3 genotype, and 36 participants with dementia onset before ETOA. (**D**) Time from meta-temporal ROI T+ to dementia (or right censored age for CDR<1), shown across ETOA and estimated amyloid burden at ETOA. Dementia was defined using total CDR scores ≥ 1; right-censored times are desaturated among participants with CDR<1. Abbreviations: A-, amyloid negative status; CDR, Clinical Dementia Rating; CL, Centiloids; ETOA, estimated tau onset age; T+, tau positive status.

### 4.4. Aim 3: Dementia trajectories with age vs. A+ time vs. T+ time

Observed CDR-SB trajectories as a function of age, A+ time, and T+ time suggested dementia onset time (i.e., CDR-SB≥4.5) had the largest range for age followed by A+ time and then T+ time (Figure 5A). With respect to age, 110/888 individuals had CDR-SB≥4.5 between ages 57 and 94 years (a 37-year range). With respect to A+ time, 102/384 A+ participants developed dementia between 0 and 25 years after A+ onset and only two remained dementia free >25 years after A+. Eighty-six of 190 T+ participants developed dementia 0 to 15 years after T+ onset. These time intervals were consistent when restricting analyses to the 176 A+T+ participants (Supplementary Figure 4).

**Figure 5.**
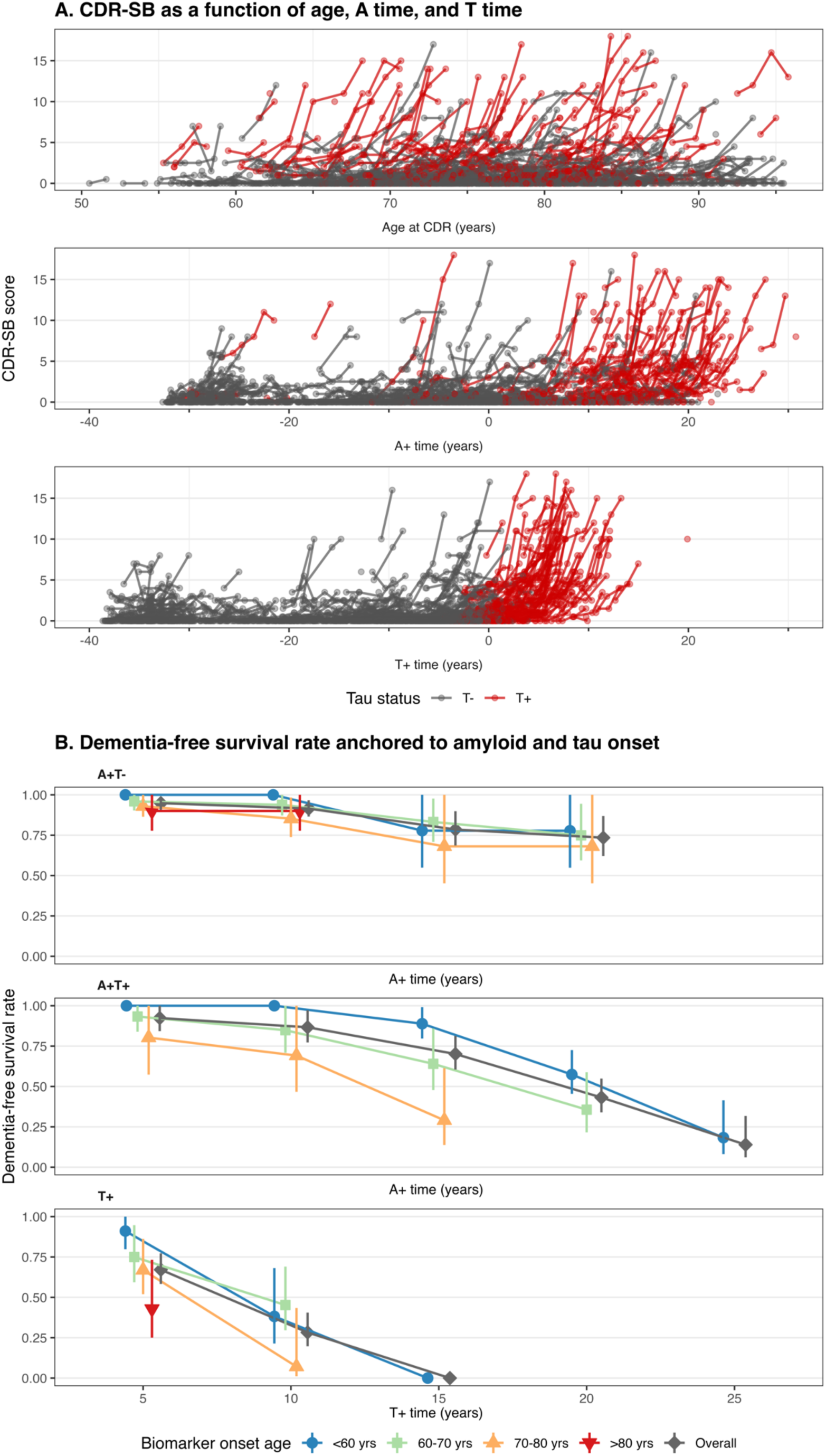
CDR-SB trajectories compared across A+ time and T+ time. (**A**) Variability in CDR-SB scores shown across chronological age (top) amyloid time (middle) and tau time (bottom). CDR-SB trajectories are highlighted in red for participants who were T+ at or before their last available tau PET scan. (**B**) 5-through 25-year dementia-free survival rates after A+ onset (n=384 A+) and after T+ onset (n=190 T+). Dementia-free probabilities were calculated using accelerated failure time models, and were stratified by decade of EAOA (top) and ETOA (bottom). Survival probabilities at 15 years were inestimable for all ETOA strata except for the group with ETOA<60 years of age. Abbreviations: A+ time, years after the onset of pathologic levels of cortical amyloid; CDR-SB, Clinical Dementia Rating Sum of Boxes score; T+ time, years after the onset of meta-temporal tau pathology.

AFT models showing dementia-free survival rates in 5-year increments after A+ and T+ onset are shown in Supplementary Figure 5. Among the A+ group (n=384), A+ time predicted a 49% (95% CI=42, 59) probability of remaining dementia-free 20 years after amyloid onset, which reduced to 16% [7, 35] probability 25 years after A+ onset. When stratifying the A+ group by T+/− (Figure 5B), 20-year dementia-free rates after A+ were 73% [62, 87] for T- and 43% [34, 55] for T+ participants, with 25-year dementia-free rates after A+ of 14% [6, 32] for T+ participants (the last T-participant converted to dementia 21.1 years after A+, rendering 25-year dementia-free rates inestimable). In contrast, dementia-free rates following T+ decreased more rapidly. In the T+ sample (n=190), the probability of remaining dementia-free was 67% [58, 77] 5 years after T+ onset, and dropped to 0% (95% CI inestimable) by 15 years after T+ onset. For both amyloid- and tau-based timelines, dementia-free rates declined faster for those with older biomarker onset age (EAOA and ETOA). Compared to the reference group with EAOA between 70-80 years of age, those who became A+ before 60 years and between 60-70 years had significantly slower time to dementia (*p*<0.001 for both). For example, 20 years after A+ onset the 70–80-year EAOA group had a 23% [9, 61] probability of remaining dementia-free, whereas the EAOA <60-year group had a 60% [48, 73] probability and the 60-70-year EAOA group had a 48% [36, 65] dementia-free probability.

For ETOA, participants with ETOA between 60-70 years had a significantly longer time from T+ to dementia compared to the ETOA 70–80 group (*p=*0.02), and those with ETOA >80 years of age had a significantly shorter time from T+ to dementia onset (*p=*0.003). For example, dementia-free survival rates 5 years after T+ were 75% [59, 95] for the ETOA 60-70 group, 67% [52, 86] for the ETOA 70–80 group and 43% [25, 73] for the ETOA >80 group. Three participants with ETOA >80 years developed dementia <5 years after T+ onset; one participant in the 70–80-year group developed dementia >10 years after T+ onset, and one participant in the <60-year group developed dementia >14 years after T+ onset. Secondary analyses calculating dementia-free survival times following T+ stratified by decade of age at baseline CDR, showed no significant age associations for amyloid and tau timelines (Supplementary Figure 6; all *p*>0.1), except for participants >80 years old at baseline who were significantly less likely to remain dementia-free after T+ onset (*p*<0.001). Repeating analyses using CDR ≥0.5 at two consecutive observations (i.e., time to MCI) indicated higher survival rate variability and no significant associations (*p*>0.1) of MCI risk with EAOA or ETOA (Supplementary Figure 7) or baseline age (Supplementary Figure 8).

## 5. Discussion

We used tau PET data from ADNI and individual-level estimated meta-temporal T+ onset ages to investigate associations of biological and selected social factors with T+ onset age and time from T+ to dementia. We contrasted these results with age and amyloid PET-based timelines and provided dementia-free survival rates vs. biomarker positive time. We found that higher amyloid at T+ onset, greater number of *APOE-ε4* alleles, and younger age at baseline tau PET were associated with younger T+ age. Furthermore, we observed that older T+ age and higher amyloid at T+ onset hastened the time from T+ to dementia. Greater literacy and higher educational attainment were associated with reduced T+ risk and later T+ onset, but these factors did not impact the time from T+ to dementia. Across analyses, sex and its interactions with *APOE* and amyloid were not significantly associated with either T+ risk or time from T+ to dementia. Overall, our findings are consistent with hypothesized and observed relationships between amyloid and tau deposition, and clinical impairment, and provide new information about the timing and interindividual variability of the timing between key AD events. This study also demonstrates how anchoring dementia onset to estimated biomarker onset age provides intuitive dementia risk characterization in terms of years of exposure. These findings have implications for clinical practice, therapeutic trials, and research investigating factors that modify AD progression.

### 5.1. Contrasting amyloid and tau PET for dementia prognosis

The revised criteria for AD diagnosis and staging supports using amyloid and tau PET for diagnosis, staging, prognosis, and as indicators for disease modifying AD therapies[1]. Our work supports this framework and provides additional context on the strengths and limitations of amyloid and tau PET for disease staging and dementia prognosis. As shown previously and replicated here with ADNI data (Figure 4A), dementia onset age in A+ people spans several decades whereas the time from A+ to dementia reduces this timeline to a ∼25-30-year window[2,5,6]. Importantly, individuals can develop dementia throughout this entire timeframe, with this variability only partially explained by established risk factors such as *APOE-ε4*, sex, and age[6]. In contrast to amyloid, our study showed only one participant remained dementia-free 15 years after T+ onset, halving the time to dementia compared to amyloid time, and only 25% remained dementia-free 10 years after T+ onset (Figure 5B). Compared to amyloid- or age-based timelines, we observed both a truncated timeline for possible dementia onset and fewer factors associated with time from T+ to dementia (Figure 5A; Supplementary Tables 4-5). Thus, when considering prognostic value, T+ onset age derived from quantitative tau PET provides substantial information about if and when dementia will occur. T+ onset age and T+ time also easily translate to clinical contexts because these time measures describe risk in years of exposure and the likelihood of dementia within a given timeframe following T+. Although A+ onset age derived from amyloid PET also has prognostic value and depicts the extended period of preclinical AD[5,6] and AD risk[43], accurate dementia prognosis with A+ onset age requires additional information such as *APOE* genotype, tau burden, and other demographics. Notably, dementia-free survival remained high (∼75%) 20 years after A+ for A+T-participants compared to ∼45% for A+T+ participants. Interestingly, higher age of biomarker positivity, but not age at baseline CDR (i.e., chronological age), was associated with greater dementia risk and shorter time from biomarker positivity to dementia. These results agree with prior work[44] and suggest that while age is a risk factor for developing AD pathology and dementia[10], once these pathologic processes are detectable with biomarkers there is likely a finite timeframe (i.e., a biomarker clock) within which individuals can remain dementia-free. This is especially true for tau deposition in AD. When considering the risk of developing AD dementia during one’s lifetime, it will be important to contrast biomarker-based risk of developing AD dementia with competing risk of death from other causes, which will aid in clinical decision making and late-life planning for people on the AD continuum. Our results also highlight the need to better characterize factors that contribute to when pathological processes begin, and factors that hasten the progression of disease from biomarker positivity to dementia.

### 5.2. Factors that affect AD progression

Although information about T+ onset age added certainty to dementia prognosis, there was still considerable heterogeneity in ETOA and dementia timing, only some of which was explained by the biological correlates of AD and SDoH we examined. As observed with EAOA[5,6] (Supplementary Figure 9), ETOA occurred earlier for *APOE-*ε4 carriers, supporting prior evidence that *APOE* modifies both amyloid and tau progression[45,46]. However, ETOA also varied by four decades within ε3ε3 and ε3ε4 genotypes. Higher amyloid burden was also related to earlier ETOA and was the only factor other than ETOA to significantly explain dementia risk following T+ onset. These findings agree with previous work suggesting the combination of amyloid and tau are strong predictors of preclinical cognitive decline and dementia[7,47,48]. While it is initially intuitive that higher amyloid burden accelerates disease progression from T+ to dementia, this relationship is also somewhat counterintuitive since higher amyloid levels at T+ onset translate to individuals having a longer period from A+ to T+ onset. A potential explanation for these effects is that people with higher amyloid at T+ onset were younger at A+ on average and were therefore likely to have fewer medical comorbidities and brain co-pathology which may impart some (relative) resistance to tau pathology. Despite a delay in the onset of tau pathology following amyloid deposition, once tau pathology is established the deleterious effects of combined amyloid and tau pathology seem insurmountable and result in relatively rapid cognitive decline and dementia. Thus, longer A+ time without tau pathology (i.e., remaining A+T-) is *de facto* protective against dementia. Despite these effects, we observed participants with ∼50 CL of amyloid converting to T+ both before 60 years and after 90 years of age, as well as participants with high amyloid and ETOA at ∼70 years convert to dementia between 2 and 12 years after T+ onset. Future work is needed to identify factors that explain interpersonal variability in A+ onset age, explicitly test variability in time from A+ onset to T+ onset, and clarify associations of these events with time to dementia onset. Additionally, although some studies suggest A+ and *APOE-*ε4 females have less resistance to tau[10,49,50] and lower cognitive resilience in the face of tau pathology, our results support other findings[44] that sex and its interactions with *APOE* and amyloid are not related to tau or dementia risk. Since *APOE*-by-sex and amyloid-by-sex cell sizes were limited, models may have been underpowered to detect interaction effects.

Beyond amyloid, *APOE,* and sex, sociodemographic associations with ETOA suggest education and literacy may influence the timing of T+ onset, and thereby dementia onset, highlighting the need to further study factors outside of biological correlates of AD pathology. Notably, these factors are modifiable. Studies widely report education effects on cognitive decline, with some suggesting that education promotes resilience at similar tau pathology levels[18,44,51–54], and others suggesting greater education delays onset of AD pathology[55]. In our study, lower education and literacy were associated with greater T+ risk and earlier T+ onset. Since time from T+ to dementia onset was predominantly impacted by ETOA, we hypothesize education and literacy (or lifestyle/environmental factors that covary with education[17,33]) could affect cognitive decline by influencing when tau deposition begins. This aligns with prior work suggesting lower education may affect early amyloid accrual and shorten the time from A+ onset to neurodegeneration and cognitive decline[55–57]. The mechanisms by which SDoH impact AD pathology must be elucidated, but it is possible that life course exposures impact the decades-long progression of AD, and psychosocial and environmental factors may have indirect effects by influencing a person’s socioeconomic and health behaviors. For instance, childhood residence in disadvantaged neighborhoods relates to faster cognitive decline by limiting educational attainment and adulthood occupation (socioeconomic mobility, intellectual stimulation)[58,59], and midlife residence amid greater air pollution is associated with higher A+ risk, neuroinflammation, and faster cognitive decline[60–64]. Although more recent research connects life course exposures with late-life cognition, many AD biomarker studies have only recently begun collecting SDoH information and often lack diversity across various SDoH[18,33,65,66]. Availability of newly collected SDoH and exposure data along with improved representation and AD biomarker characterization will enable more thorough investigation of how life course environmental and social determinants potentially influence AD pathophysiology and AD-related dementia risk.

### 5.3. Strengths and Limitations

One strength of this study is the ability of temporal modeling to estimate when amyloid and tau biomarkers become abnormal. This enables investigation of factors that influence AD biomarker onset and time from biomarker onset to dementia without directly observing key biomarker events. Among the limitations were limited representation among non-White/Hispanic individuals and participants with high school education or less, which may limit the generalizability of these findings. Future studies with better representation in these and other domains are needed to determine the extent to which these findings will generalize across population groups.

## Abbreviations

AD: Alzheimer’s disease
Aβ: amyloid β
AD dementia: dementia suspected due to AD
ADNI: Alzheimer’s Disease Neuroimaging Initiative
*APOE*: Apolipoprotein E
CDR: Clinical Dementia Rating
CDR-SB: Clinical Dementia Rating – Sum of Boxes
CL: Centiloids
CSF: cerebrospinal fluid
CU: cognitively unimpaired
FBB: ^18^F-Florbetaben
FBP: ^18^F-Florbetapir
FTP: ^18^F-Flortaucipir
GMM: Gaussian mixture modeling
HR: hazard ratio
MCI: mild cognitive impairment
MRI: magnetic resonance imaging
PET: positron emission tomography
ptau181: phosphorylated tau 181
SDoH: social determinants of health
SILA: sampled iterative local approximation
SUVR: standardized uptake value ratio

## Acknowledgments

Data collection and sharing for this project was funded by the Alzheimer’s Disease Neuroimaging Initiative (ADNI) (National Institutes of Health Grant U01 AG024904) and DOD ADNI (Department of Defense award number W81XWH-12-2-0012). ADNI is funded by the National Institute on Aging, the National Institute of Biomedical Imaging and Bioengineering, and through generous contributions from the following: AbbVie, Alzheimer’s Association; Alzheimer’s Drug Discovery Foundation; Araclon Biotech; BioClinica, Inc.; Biogen; Bristol-Myers Squibb Company; CereSpir, Inc.; Cogstate; Eisai Inc.; Elan Pharmaceuticals, Inc.; Eli Lilly and Company; EuroImmun; F. Hoffmann-La Roche Ltd and its affiliated company Genentech, Inc.; Fujirebio; GE Healthcare; IXICO Ltd.; Janssen Alzheimer Immunotherapy Research & Development, LLC.; Johnson & Johnson Pharmaceutical Research & Development LLC.; Lumosity; Lundbeck; Merck & Co., Inc.; Meso Scale Diagnostics, LLC.; NeuroRx Research; Neurotrack Technologies; Novartis Pharmaceuticals Corporation; Pfizer Inc.; Piramal Imaging; Servier; Takeda Pharmaceutical Company; and Transition Therapeutics. The Canadian Institutes of Health Research is providing funds to support ADNI clinical sites in Canada. Private sector contributions are facilitated by the Foundation for the National Institutes of Health (www.fnih.org). The grantee organization is the Northern California Institute for Research and Education, and the study is coordinated by the Alzheimer’s Therapeutic Research Institute at the University of Southern California. ADNI data are disseminated by the Laboratory for Neuro Imaging at the University of Southern California. For CEG, this material is the result of work supported with resources and the use of facilities at the William S. Middleton Memorial VA Hospital in Madison, Wisconsin.

## Consent Statement

Written consent was obtained before enrollment in accordance with the Declaration of Helsinki standards and under Institutional Review Board review for each source study, including the University of Wisconsin Institutional Review Board.

## References

[1] Jack CR, Andrews JS, Beach TG, Buracchio T, Dunn B, Graf A, et al. Revised criteria for diagnosis and staging of Alzheimer’s disease: Alzheimer’s Association Workgroup. Alzheimer’s & Dementia 2024:alz.13859. 10.1002/alz.13859.

[2] Villemagne VL, Burnham S, Bourgeat P, Brown B, Ellis KA, Salvado O, et al. Amyloid β deposition, neurodegeneration, and cognitive decline in sporadic Alzheimer’s disease: a prospective cohort study. The Lancet Neurology 2013;12:357–67. 10.1016/S1474-4422(13)70044-9.

[3] Bilgel M, An Y, Zhou Y, Wong DF, Prince JL, Ferrucci L, et al. Individual estimates of age at detectable amyloid onset for risk factor assessment. Alzheimers Dement 2016;12:373–9. 10.1016/j.jalz.2015.08.166.

[4] Insel PS, Donohue MC, Berron D, Hansson O, Mattsson-Carlgren N. Time between milestone events in the Alzheimer’s disease amyloid cascade. Neuroimage 2021;227:117676. 10.1016/j.neuroimage.2020.117676.

[5] Schindler SE, Li Y, Buckles VD, Gordon BA, Benzinger TLS, Wang G, et al. Predicting Symptom Onset in Sporadic Alzheimer Disease With Amyloid PET. Neurology 2021;97:e1823–34. 10.1212/WNL.0000000000012775.

[6] Betthauser TJ, Bilgel M, Koscik RL, Jedynak BM, An Y, Kellett KA, et al. Multi-method investigation of factors influencing amyloid onset and impairment in three cohorts. Brain 2022;145:4065–79. 10.1093/brain/awac213.

[7] Ossenkoppele R, Pichet Binette A, Groot C, Smith R, Strandberg O, Palmqvist S, et al. Amyloid and tau PET-positive cognitively unimpaired individuals are at high risk for future cognitive decline. Nat Med 2022;28:2381–7. 10.1038/s41591-022-02049-x.

[8] Barthélemy NR, Li Y, Joseph-Mathurin N, Gordon BA, Hassenstab J, Benzinger TLS, et al. A soluble phosphorylated tau signature links tau, amyloid and the evolution of stages of dominantly inherited Alzheimer’s disease. Nat Med 2020;26:398–407. 10.1038/s41591-020-0781-z.

[9] Salvadó G, Horie K, Barthélemy NR, Vogel JW, Pichet Binette A, Chen CD, et al. Disease staging of Alzheimer’s disease using a CSF-based biomarker model. Nat Aging 2024;4:694–708. 10.1038/s43587-024-00599-y.

[10] Arenaza-Urquijo EM, Boyle R, Casaletto K, Anstey KJ, Vila-Castelar C, Colverson A, et al. Sex and gender differences in cognitive resilience to aging and Alzheimer’s disease. Alzheimer’s & Dementia 2024:alz.13844. 10.1002/alz.13844.

[11] Buckley RF, Mormino EC, Rabin JS, Hohman TJ, Landau S, Hanseeuw BJ, et al. Sex Differences in the Association of Global Amyloid and Regional Tau Deposition Measured by Positron Emission Tomography in Clinically Normal Older Adults. JAMA Neurol 2019;76:542. 10.1001/jamaneurol.2018.4693.

[12] Buckley RF, Scott MR, Jacobs HIL, Schultz AP, Properzi MJ, Amariglio RE, et al. Sex Mediates Relationships Between Regional Tau Pathology and Cognitive Decline. Ann Neurol 2020;88:921–32. 10.1002/ana.25878.

[13] Altmann A, Tian L, Henderson VW, Greicius MD, Investigators ADNI. Sex modifies the APOE-related risk of developing Alzheimer disease. Annals of Neurology 2014;75:563–73. 10.1002/ana.24135.

[14] Hohman TJ, Dumitrescu L, Barnes LL, Thambisetty M, Beecham G, Kunkle B, et al. Sex-Specific Association of Apolipoprotein E With Cerebrospinal Fluid Levels of Tau. JAMA Neurol 2018;75:989. 10.1001/jamaneurol.2018.0821.

[15] Buckley RF, Mormino EC, Chhatwal J, Schultz AP, Rabin JS, Rentz DM, et al. Associations between baseline amyloid, sex, and APOE on subsequent tau accumulation in cerebrospinal fluid. Neurobiol Aging 2019;78:178–85. 10.1016/j.neurobiolaging.2019.02.019.

[16] Zuelsdorff M, Okonkwo OC, Norton D, Barnes LL, Graham KL, Clark LR, et al. Stressful Life Events and Racial Disparities in Cognition Among Middle-Aged and Older Adults. JAD 2020;73:671–82. 10.3233/JAD-190439.

[17] Adkins-Jackson PB, George KM, Besser LM, Hyun J, Lamar M, Hill-Jarrett TG, et al. The structural and social determinants of Alzheimer’s disease related dementias. Alzheimer’s & Dementia 2023;19:3171–85. 10.1002/alz.13027.

[18] Livingston G, Huntley J, Liu KY, Costafreda SG, Selbæk G, Alladi S, et al. Dementia prevention, intervention, and care: 2024 report of the Lancet standing Commission. The Lancet 2024;404:572–628. 10.1016/S0140-6736(24)01296-0.

[19] Hunt JFV, Buckingham W, Kim AJ, Oh J, Vogt NM, Jonaitis EM, et al. Association of Neighborhood-Level Disadvantage With Cerebral and Hippocampal Volume. JAMA Neurol 2020;77:451. 10.1001/jamaneurol.2019.4501.

[20] Hunt JFV, Vogt NM, Jonaitis EM, Buckingham WR, Koscik RL, Zuelsdorff M, et al. Association of Neighborhood Context, Cognitive Decline, and Cortical Change in an Unimpaired Cohort. Neurology 2021;96:e2500–12. 10.1212/WNL.0000000000011918.

[21] Ramanan VK, Castillo AM, Knopman DS, Graff-Radford J, Lowe VJ, Petersen RC, et al. Association of Apolipoprotein E ɛ4, Educational Level, and Sex With Tau Deposition and Tau-Mediated Metabolic Dysfunction in Older Adults. JAMA Netw Open 2019;2:e1913909. 10.1001/jamanetworkopen.2019.13909.

[22] Koscik RL, Betthauser TJ, Jonaitis EM, Allison SL, Clark LR, Hermann BP, et al. Amyloid duration is associated with preclinical cognitive decline and tau PET. Alzheimer’s & Dementia: Diagnosis, Assessment & Disease Monitoring 2020;12. 10.1002/dad2.12007.

[23] Ashford MT, Raman R, Miller G, Donohue MC, Okonkwo OC, Mindt MR, et al. Screening and enrollment of underrepresented ethnocultural and educational populations in the Alzheimer’s Disease Neuroimaging Initiative (ADNI). Alzheimer’s & Dementia 2022;18:2603–13. 10.1002/alz.12640.

[24] Mindt MR, Okonkwo O, Weiner MW, Veitch DP, Aisen P, Ashford M, et al. Improving generalizability and study design of Alzheimer’s disease cohort studies in the United States by including under-represented populations. Alzheimer’s & Dementia 2023;19:1549–57. 10.1002/alz.12823.

[25] Veitch DP, Weiner MW, Miller M, Aisen PS, Ashford MA, Beckett LA, et al. The Alzheimer’s Disease Neuroimaging Initiative in the era of Alzheimer’s disease treatment: A review of ADNI studies from 2021 to 2022. Alzheimer’s & Dementia 2024;20:652–94. 10.1002/alz.13449.

[26] Trojanowski JQ, Vandeerstichele H, Korecka M, Clark CM, Aisen PS, Petersen RC, et al. Update on the biomarker core of the Alzheimer’s Disease Neuroimaging Initiative subjects. Alzheimer’s & Dementia 2010;6:230–8. 10.1016/j.jalz.2010.03.008.

[27] Petersen RC, Aisen PS, Beckett LA, Donohue MC, Gamst AC, Harvey DJ, et al. Alzheimer’s Disease Neuroimaging Initiative (ADNI): Clinical characterization. Neurology 2010;74:201–9. 10.1212/WNL.0b013e3181cb3e25.

[28] McKhann G, Drachman D, Folstein M, Katzman R, Price D, Stadlan EM. Clinical diagnosis of Alzheimer’s disease: Report of the NINCDS-ADRDA Work Group* under the auspices of Department of Health and Human Services Task Force on Alzheimer’s Disease. Neurology 1984;34:939–939. 10.1212/WNL.34.7.939.

[29] Morris JC. The Clinical Dementia Rating (CDR): current version and scoring rules. Neurology 1993;43:2412–4. 10.1212/wnl.43.11.2412-a.

[30] O’Bryant SE. Staging Dementia Using Clinical Dementia Rating Scale Sum of Boxes Scores: A Texas Alzheimer’s Research Consortium Study. Arch Neurol 2008;65:1091. 10.1001/archneur.65.8.1091.

[31] Sisco S, Gross AL, Shih RA, Sachs BC, Glymour MM, Bangen KJ, et al. The role of early-life educational quality and literacy in explaining racial disparities in cognition in late life. J Gerontol B Psychol Sci Soc Sci 2015;70:557–67. 10.1093/geronb/gbt133.

[32] Sharp ES, Gatz M. Relationship Between Education and Dementia: An Updated Systematic Review. Alzheimer Disease & Associated Disorders 2011;25:289–304. 10.1097/WAD.0b013e318211c83c.

[33] Stites SD, Midgett S, Mechanic-Hamilton D, Zuelsdorff M, Glover CM, Marquez DX, et al. Establishing a Framework for Gathering Structural and Social Determinants of Health in Alzheimer’s Disease Research Centers. The Gerontologist 2022;62:694–703. 10.1093/geront/gnab182.

[34] Grober E, Sliwinski M. Development and validation of a model for estimating premorbid verbal intelligence in the elderly. J Clin Exp Neuropsychol 1991;13:933–49. 10.1080/01688639108405109.

[35] Fromm D, Holland AL, Nebes RD, Oakley MA. A longitudinal study of word-reading ability in Alzheimer’s disease: evidence from the National Adult Reading Test. Cortex 1991;27:367–76. 10.1016/s0010-9452(13)80032-9.

[36] Rentz DM, Locascio JJ, Becker JA, Moran EK, Eng E, Buckner RL, et al. Cognition, reserve, and amyloid deposition in normal aging. Ann Neurol 2010;67:353–64. 10.1002/ana.21904.

[37] Paolo AM, Ryan JJ. Generalizability of two methods of estimating premorbid intelligence in the elderly. Arch Clin Neuropsychol 1992;7:135–43.

[38] Jagust WJ, Landau SM, Shaw LM, Trojanowski JQ, Koeppe RA, Reiman EM, et al. Relationships between biomarkers in aging and dementia. Neurology 2009;73:1193–9. 10.1212/WNL.0b013e3181bc010c.

[39] Mormino EC, Kluth JT, Madison CM, Rabinovici GD, Baker SL, Miller BL, et al. Episodic memory loss is related to hippocampal-mediated β-amyloid deposition in elderly subjects. Brain 2009;132:1310–23. 10.1093/brain/awn320.

[40] Landau SM, Fero A, Baker SL, Koeppe R, Mintun M, Chen K, et al. Measurement of longitudinal β-amyloid change with 18F-florbetapir PET and standardized uptake value ratios. J Nucl Med 2015;56:567–74. 10.2967/jnumed.114.148981.

[41] for the Alzheimer’s Disease Neuroimaging Initiative, Royse SK, Minhas DS, Lopresti BJ, Murphy A, Ward T, et al. Validation of amyloid PET positivity thresholds in centiloids: a multisite PET study approach. Alz Res Therapy 2021;13:99. 10.1186/s13195-021-00836-1.

[42] Kassambara A, Kosinski M, Biecek P. survminer: Drawing Survival Curves using “ggplot2” 2016:0.4.9. 10.32614/CRAN.package.survminer.

[43] Dubois B, Villain N, Schneider L, Fox N, Campbell N, Galasko D, et al. Alzheimer Disease as a Clinical-Biological Construct—An International Working Group Recommendation. JAMA Neurology 2024;81:1304–11. 10.1001/jamaneurol.2024.3770.

[44] Bocancea DI, Svenningsson AL, Van Loenhoud AC, Groot C, Barkhof F, Strandberg O, et al. Determinants of cognitive and brain resilience to tau pathology: a longitudinal analysis. Brain 2023;146:3719–34. 10.1093/brain/awad100.

[45] Neitzel J, Franzmeier N, Rubinski A, Binette AP, Poirier J, Villeneuve S, et al. ApoE4 associated with higher tau accumulation independent of amyloid burden: Human neuropathology/tau. Alzheimer’s & Dementia 2020;16:e046206. 10.1002/alz.046206.

[46] Baek MS, Cho H, Lee HS, Lee JH, Ryu YH, Lyoo CH. Effect of APOE ε4 genotype on amyloid-β and tau accumulation in Alzheimer’s disease. Alz Res Therapy 2020;12:140. 10.1186/s13195-020-00710-6.

[47] Brier MR, Gordon B, Friedrichsen K, McCarthy J, Stern A, Christensen J, et al. Tau and Aβ imaging, CSF measures, and cognition in Alzheimer’s disease. Sci Transl Med 2016;8:338ra66. 10.1126/scitranslmed.aaf2362.

[48] Jack CR, Wiste HJ, Botha H, Weigand SD, Therneau TM, Knopman DS, et al. The bivariate distribution of amyloid-β and tau: relationship with established neurocognitive clinical syndromes. Brain 2019;142:3230–42. 10.1093/brain/awz268.

[49] Coughlan GT, Betthauser TJ, Boyle R, Koscik RL, Klinger HM, Chibnik LB, et al. Association of Age at Menopause and Hormone Therapy Use With Tau and β-Amyloid Positron Emission Tomography. JAMA Neurol 2023;80:462–73. 10.1001/jamaneurol.2023.0455.

[50] Wang Y-TT, Pascoal TA, Therriault J, Kang MS, Benedet AL, Savard M, et al. Interactive rather than independent effect of APOE and sex potentiates tau deposition in women. Brain Commun 2021;3:fcab126. 10.1093/braincomms/fcab126.

[51] Bauer CE, Brown CA, Gold BT, Alzheimer’s Disease Neuroimaging Initiative. Education does not protect cognitive function from brain pathology in the ADNI 2 cohort. Neurobiol Aging 2020;90:147–9. 10.1016/j.neurobiolaging.2019.11.017.

[52] Rentz DM, Mormino EC, Papp KV, Betensky RA, Sperling RA, Johnson KA. Cognitive resilience in clinical and preclinical Alzheimer’s disease: the Association of Amyloid and Tau Burden on cognitive performance. Brain Imaging Behav 2017;11:383–90. 10.1007/s11682-016-9640-4.

[53] Hoenig MC, Bischof GN, Hammes J, Faber J, Fliessbach K, van Eimeren T, et al. Tau pathology and cognitive reserve in Alzheimer’s disease. Neurobiology of Aging 2017;57:1–10.1016/j.neurobiolaging.2017.05.004.

[54] Hoenig MC, Bischof GN, Onur ÖA, Kukolja J, Jessen F, Fliessbach K, et al. Level of education mitigates the impact of tau pathology on neuronal function. Eur J Nucl Med Mol Imaging 2019;46:1787–95. 10.1007/s00259-019-04342-3.

[55] Arenaza-Urquijo EM, Bejanin A, Gonneaud J, Wirth M, La Joie R, Mutlu J, et al. Association between educational attainment and amyloid deposition across the spectrum from normal cognition to dementia: neuroimaging evidence for protection and compensation. Neurobiology of Aging 2017;59:72–9. 10.1016/j.neurobiolaging.2017.06.016.

[56] Vemuri P, Weigand SD, Przybelski SA, Knopman DS, Smith GE, Trojanowski JQ, et al. Cognitive reserve and Alzheimer’s disease biomarkers are independent determinants of cognition. Brain 2011;134:1479. 10.1093/brain/awr049.

[57] Lo RY, Jagust WJ, Alzheimer’s Disease Neuroimaging Initiative. Effect of cognitive reserve markers on Alzheimer pathologic progression. Alzheimer Dis Assoc Disord 2013;27:343–50. 10.1097/WAD.0b013e3182900b2b.

[58] Baranyi G, Conte F, Deary IJ, Shortt N, Thompson CW, Cox SR, et al. Neighbourhood deprivation across eight decades and late-life cognitive function in the Lothian Birth Cohort 1936: a life-course study. Age Ageing 2023;52:afad056. 10.1093/ageing/afad056.

[59] Marden JR, Tchetgen Tchetgen EJ, Kawachi I, Glymour MM. Contribution of Socioeconomic Status at 3 Life-Course Periods to Late-Life Memory Function and Decline: Early and Late Predictors of Dementia Risk. American Journal of Epidemiology 2017;186:805–14. 10.1093/aje/kwx155.

[60] Iaccarino L, La Joie R, Lesman-Segev OH, Lee E, Hanna L, Allen IE, et al. Association Between Ambient Air Pollution and Amyloid Positron Emission Tomography Positivity in Older Adults With Cognitive Impairment. JAMA Neurol 2021;78:197. 10.1001/jamaneurol.2020.3962.

[61] Lee Y, Yoon S, Yoon SH, Kang SW, Jeon S, Kim M, et al. Air pollution is associated with faster cognitive decline in Alzheimer’s disease. Ann Clin Transl Neurol 2023;10:964–73. 10.1002/acn3.51779.

[62] Levesque S, Surace MJ, McDonald J, Block ML. Air pollution & the brain: Subchronic diesel exhaust exposure causes neuroinflammation and elevates early markers of neurodegenerative disease. J Neuroinflammation 2011;8:105. 10.1186/1742-2094-8-105.

[63] Casey E, Li Z, Liang D, Ebelt S, Levey AI, Lah JJ, et al. Association between Fine Particulate Matter Exposure and Cerebrospinal Fluid Biomarkers of Alzheimer’s Disease among a Cognitively Healthy Population-Based Cohort. Environ Health Perspect 2024;132:47001. 10.1289/EHP13503.

[64] Ma Y-H, Chen H-S, Liu C, Feng Q-S, Feng L, Zhang Y-R, et al. Association of Long-term Exposure to Ambient Air Pollution With Cognitive Decline and Alzheimer’s Disease– Related Amyloidosis. Biological Psychiatry 2023;93:780–9. 10.1016/j.biopsych.2022.05.017.

[65] Livingston G, Huntley J, Sommerlad A, Ames D, Ballard C, Banerjee S, et al. Dementia prevention, intervention, and care: 2020 report of the Lancet Commission. The Lancet 2020;396:413–46. 10.1016/S0140-6736(20)30367-6.

[66] Bennett DA, Arnold SE, Valenzuela MJ, Brayne C, Schneider JA. Cognitive and social lifestyle: links with neuropathology and cognition in late life. Acta Neuropathol 2014;127:137–50. 10.1007/s00401-013-1226-2.

