## Supplemental Materials for "Factors associated with age at tau pathology onset and time from tau onset to dementia"

**Supplementary Materials**

**Supplemental Figures**

### A. $^{18}$F-Florbetapir and $^{18}$F-Florbetaben cortical ROI

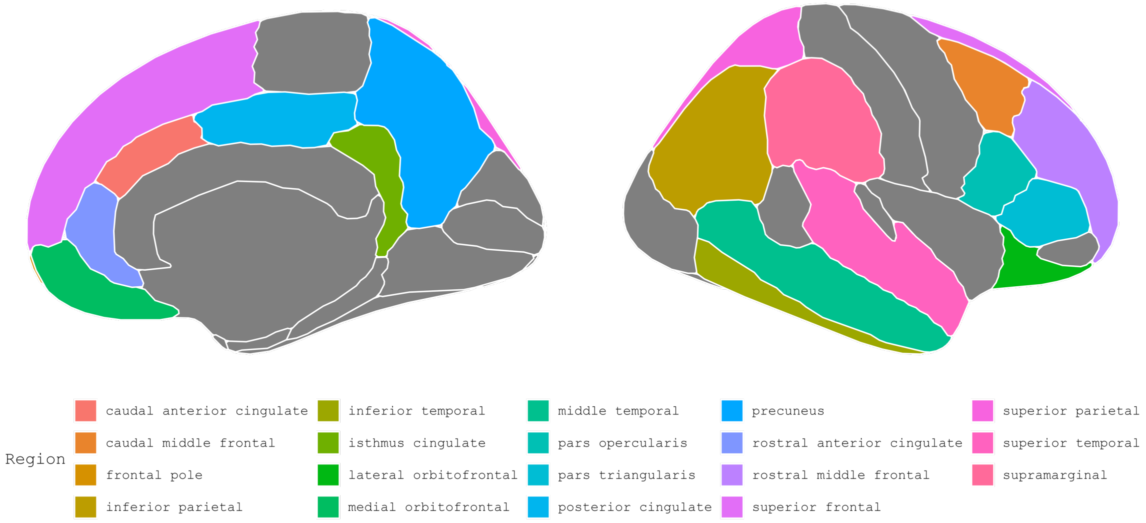


### B. $^{18}$F-Flortaucipir Meta-temporal ROI

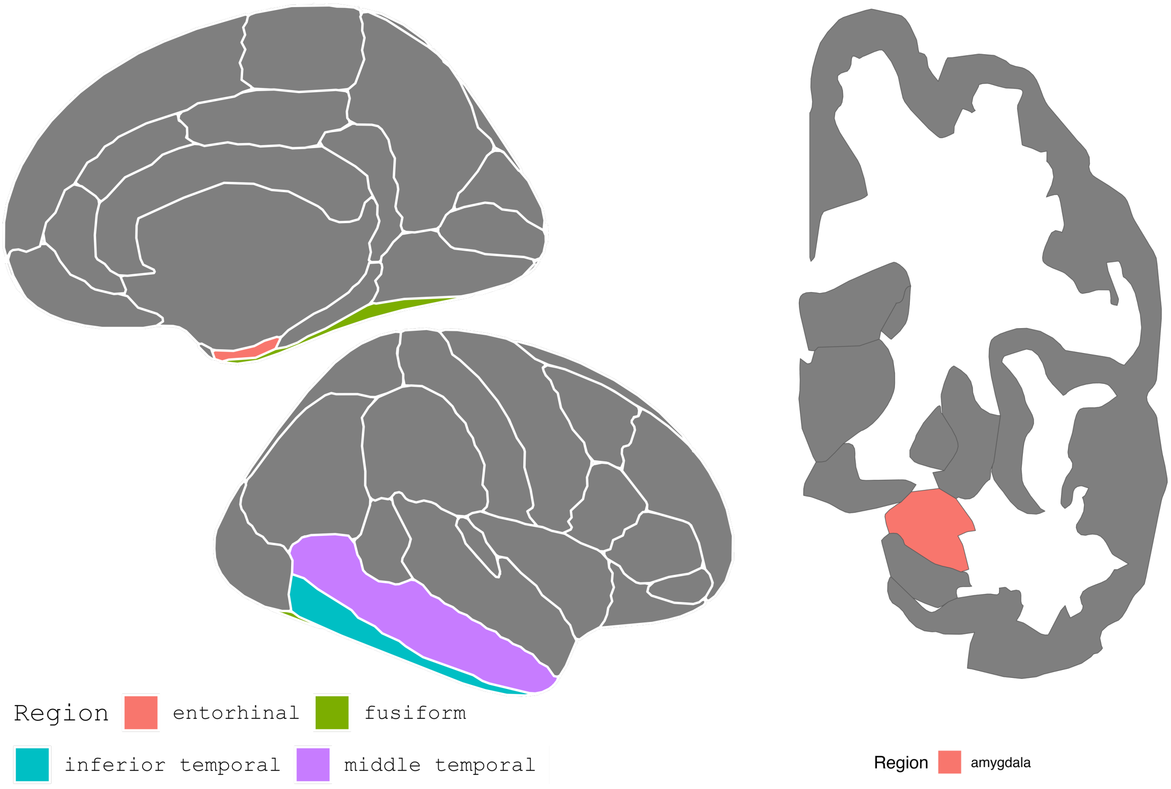

**Supplemental Figure 1. Regions of interest used to quantify amyloid and tau burden.** ROIs were defined bilaterally; right hemisphere is shown for demonstrative purposes. (**A**) 19 cortical ROIs from which $^{18}$F-Florbetapir and $^{18}$F-Florbetaben SUVR was extracted to quantify amyloid burden. (**B**) 5 regions comprising the meta-temporal ROI from which $^{18}$F-Flortaucipir SUVR was extracted to quantify tau burden. SUVRs were quantified by the University of California, Berkeley PET core[1]; derived values were converted to Centiloids[2] and used in analysis for the present study. Abbreviations: PET, positron emission tomography; ROI, region of interest; SUVR, standardized uptake value ratio.

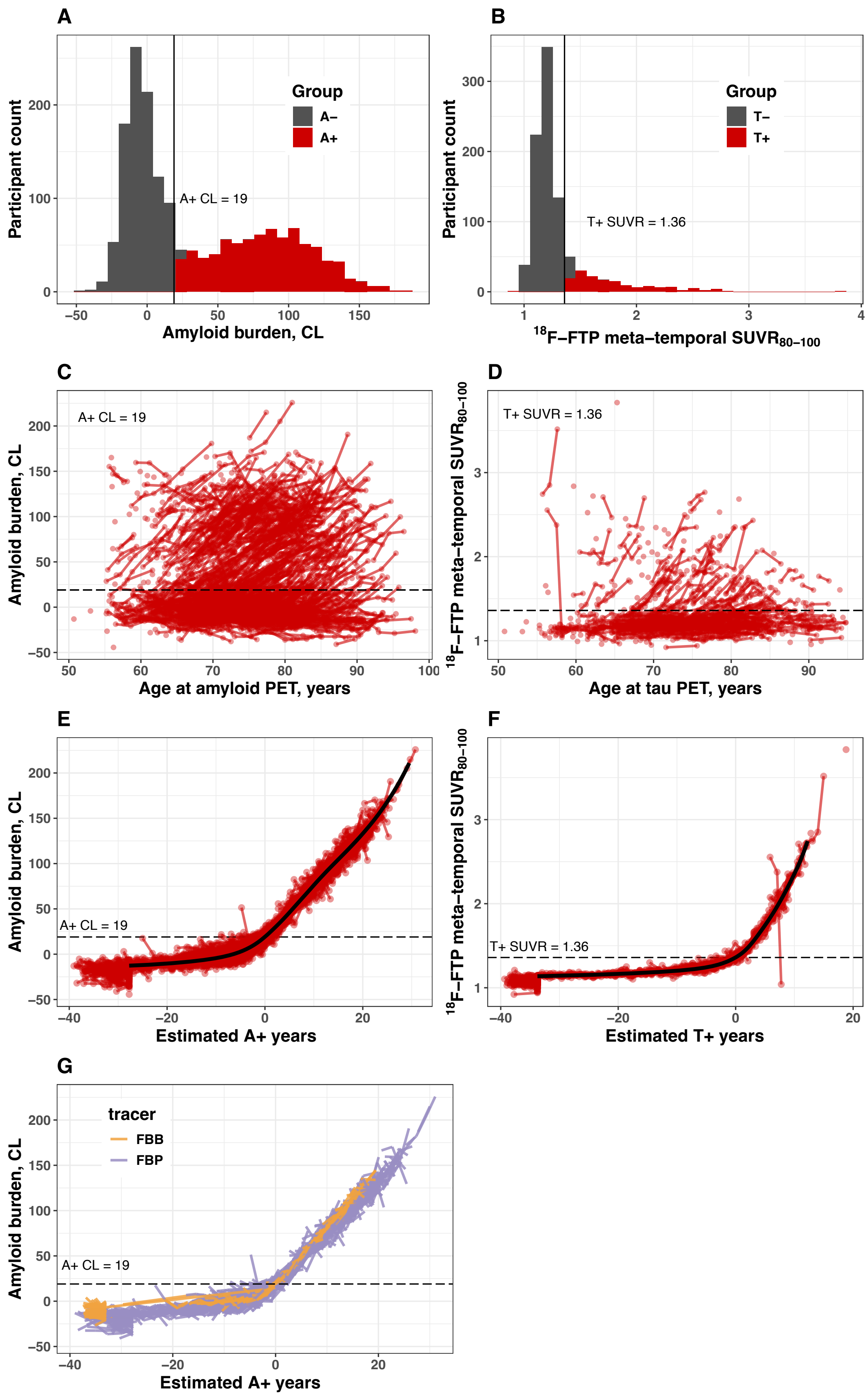

A
Participant count
Amyloid burden, CL
Group
A–
A+
A+ CL = 19
B
$^{18}$F–FTP meta–temporal SUVR$_{80–100}$
T–
T+
T+ SUVR = 1.36
C
Amyloid burden, CL
Age at amyloid PET, years
D
$^{18}$F–FTP meta–temporal SUVR$_{80–100}$
Age at tau PET, years
E
Estimated A+ years
F
Estimated T+ years
G
tracer
FBB
FBP

**Supplemental Figure 2. Biomarker positivity and SILA modeling for amyloid and tau PET data.** Modeling steps are depicted for amyloid on the left and tau on the right. (**A-B**) Gaussian mixture modeling used to classify biomarker negative and positive participants; positivity was two standard deviations above the mean of the lower group. Horizontal dotted lines in (**C-G**) indicate the derived positivity thresholds. (**C-D**) Spaghetti plots showing cross-sectional and longitudinal PET observations for every ADNI participant with either amyloid or tau data. (**E-F**) SILA-modeled mean biomarker trajectory (black line) and estimated biomarker trajectory per participant (red lines). Every participant with either amyloid or tau data was included to obtain robust models of each biomarker trajectory; participants with both amyloid and tau data were subsequently selected for inclusion in statistical analysis. (**G**) SILA-estimated amyloid trajectories modeled separately for FBB and FBP. While slopes differ by approximately 1 CL/year between tracers, given that A+ individuals accumulate amyloid at a rate of 5 CL/year, this difference in slopes results in negligible differences in biomarker time estimates (0.20 year). Consequently, tracers were considered to have strong concordance in their SILA models, and a jointly modeled SILA trajectory (**E**) was used for statistical analysis. Abbreviations: A, amyloid positive (+) or negative (-) status; CL, Centiloids; FBB, $^{18}$F-Florbetaben; FBP, $^{18}$F-Florbetapir; PET, FTP, $^{18}$F-Flortaucipir; PET, positron emission tomography; SILA, sampled iterative localized approximation; SUVR, standardized uptake value ratio; T, tau positive (+) or negative (-) status.

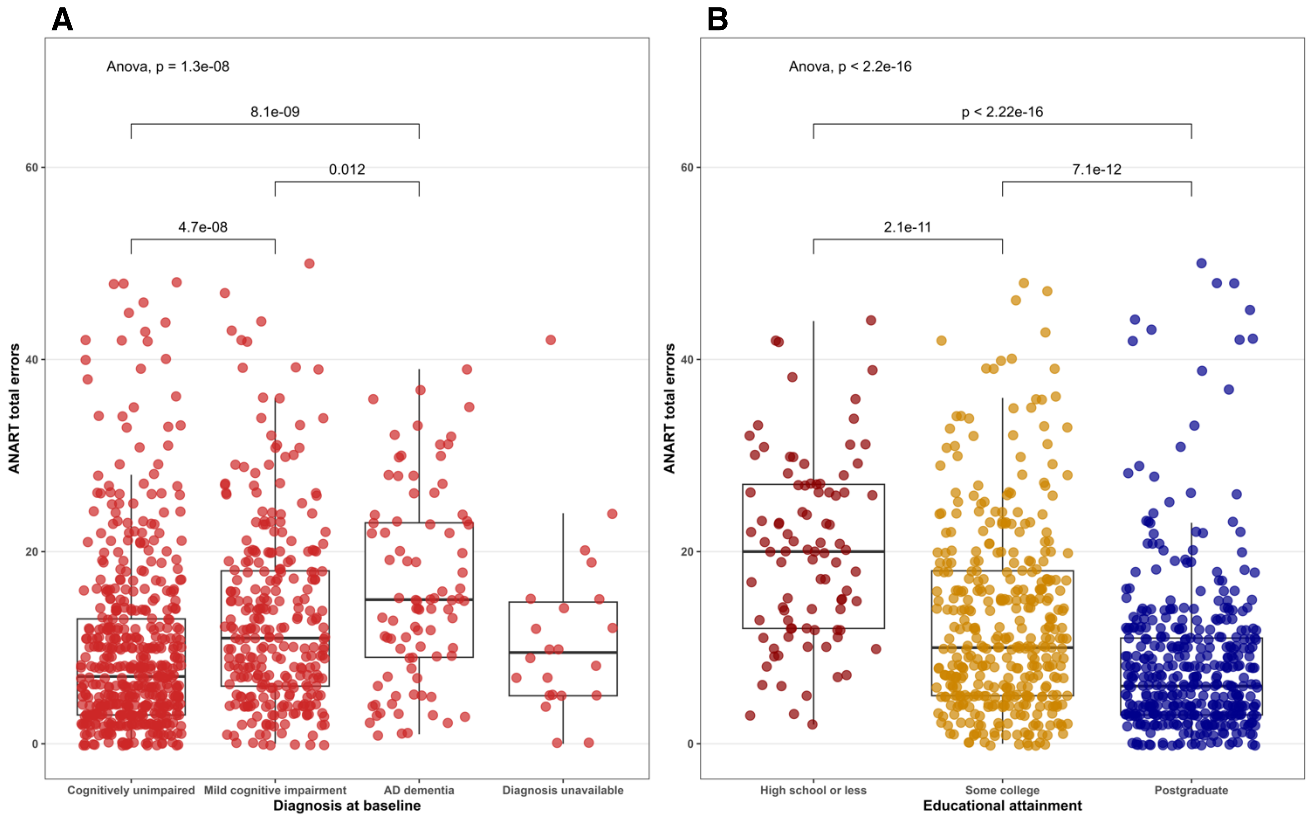


**Supplemental Figure 3. ANART total errors distributions across diagnosis and educational attainment.** Total reading errors on the ANART plotted for the full sample with amyloid and tau PET data (N=905) across clinical diagnosis at baseline visit (**A**) and self-reported educational attainment (**B**). Differences in mean reading errors were tested across conditions using one-way ANOVA. Because reading error rates differed significantly across the full cohort, pairwise comparisons of reading errors across groups were assessed using t-tests. In (**A**), significance testing excluded participants without clinical diagnosis data. Abbreviations: AD, Alzheimer's disease; ANART, American National Adult Reading Test; ANOVA, analysis of variance.

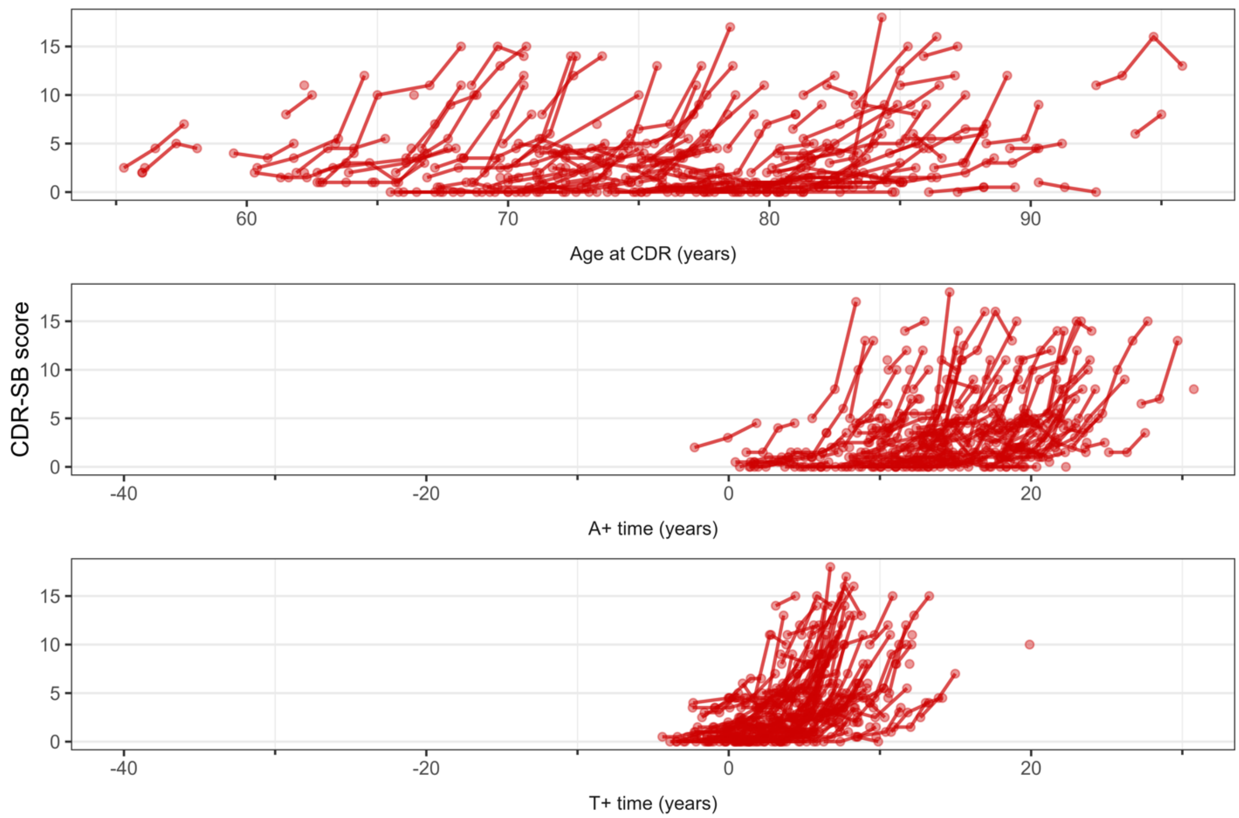


**Supplemental Figure 4. CDR-SB trajectories compared across A+ time and T+ time in the A+T+ subset.** Variability in CDR-SB scores shown across chronological age (top) amyloid time (middle) and tau time (bottom) for participants with A+T+ biomarker status (n=176). Abbreviations: A+ time, estimated years A+ from SILA; CDR-SB, Clinical Dementia Rating Sum of Boxes score; T+ time, estimated years T+ in the meta-temporal ROI from SILA.

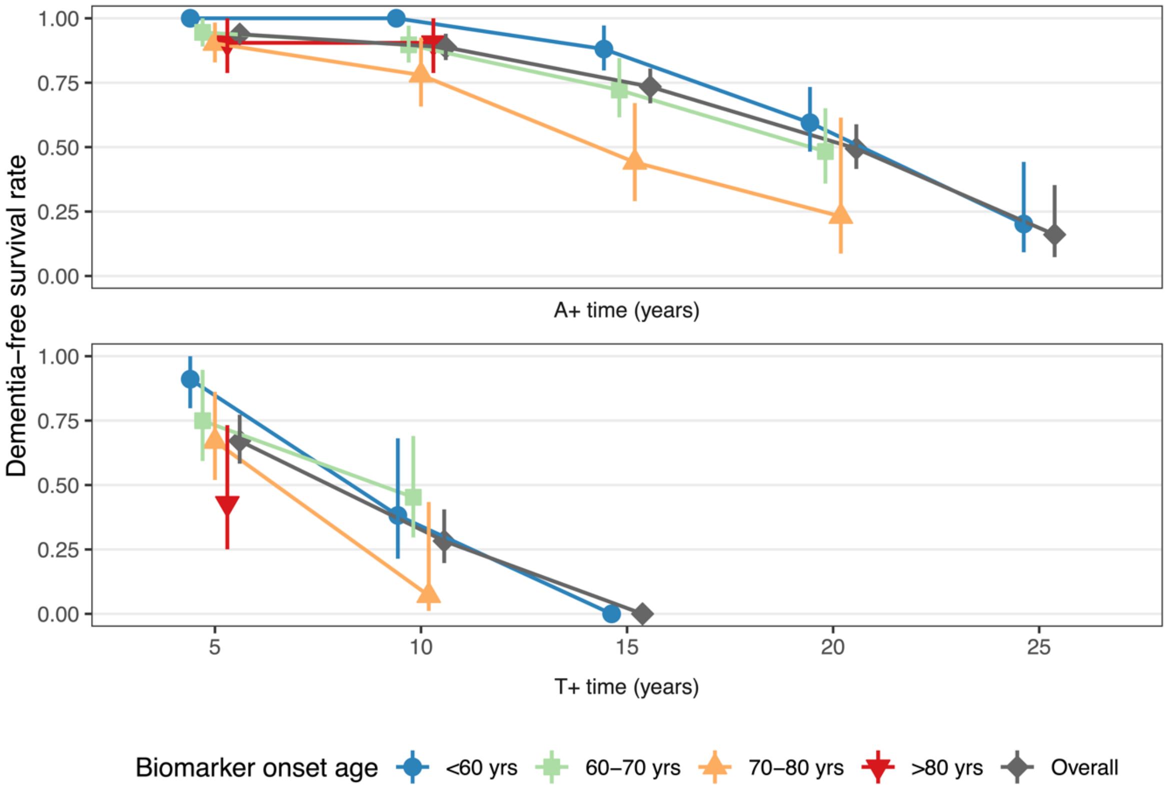


**Supplemental Figure 5. Dementia-free survival rate anchored to A+ and meta-temporal T+ onset.** 5- through 25-year dementia-free survival rates after A+ onset (n=384 A+) and after T+ onset (n=190 T+). Dementia-free probabilities were calculated using accelerated failure time models and were stratified by decade of EAOA (top) and ETOA (bottom). Survival probabilities at 15 years were inestimable for all ETOA strata except for the group with ETOA<60 years of age. Abbreviations: A+ time, estimated years A+ from SILA; T+ time, estimated years T+ in the meta-temporal ROI from SILA.

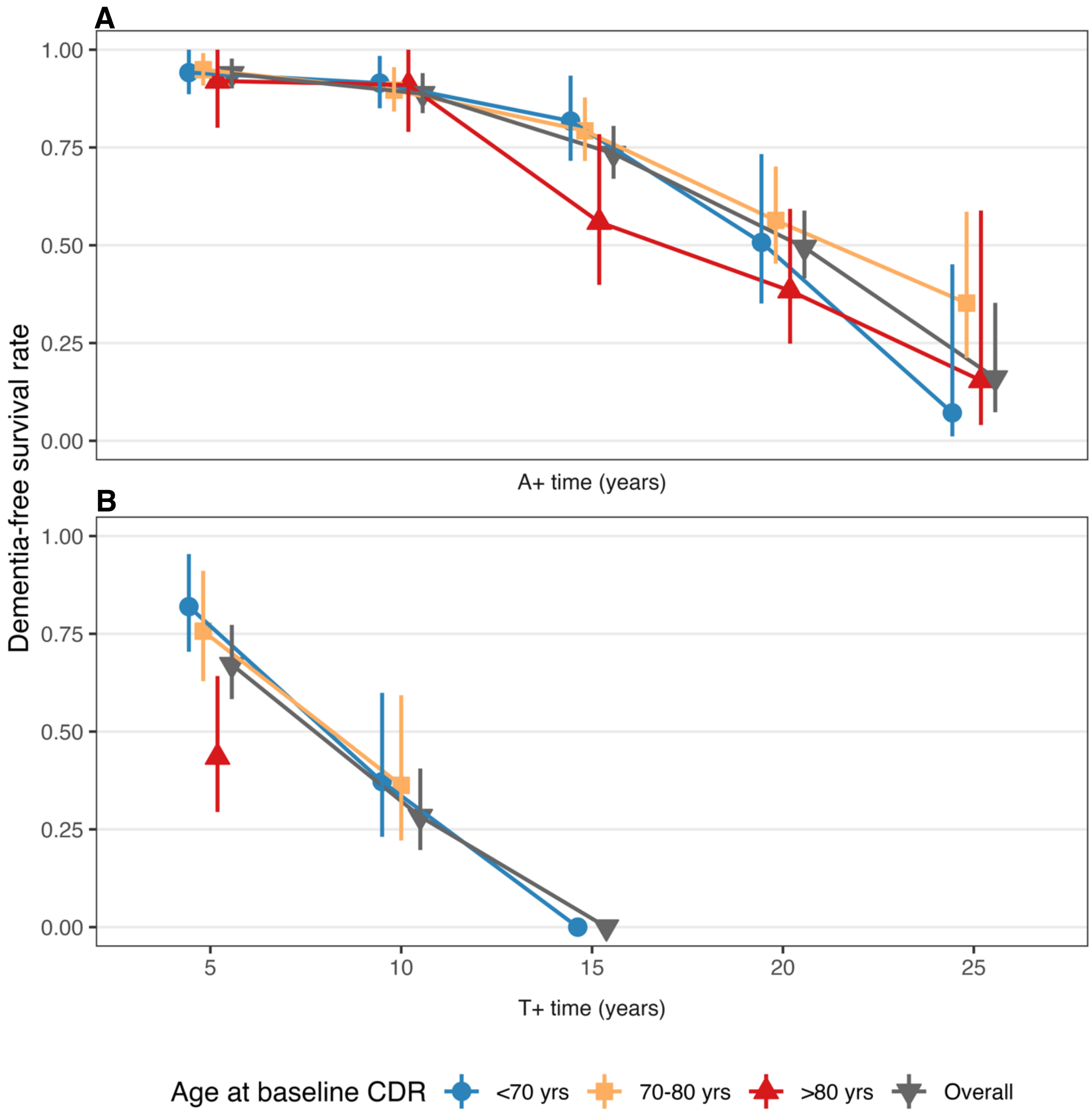


**Supplemental Figure 6. Dementia-free survival rates across A+ time and T+ time, stratified by age at baseline CDR.** Individuals were considered to have dementia if they had a CDR global score of one or greater. 5- through 25-year dementia-free survival rates after (**A**) A+ onset (n=384 A+) and (**B**) T+ onset (n=190 T+). Dementia-free probabilities were calculated using accelerated failure time models, and were stratified by decade of baseline CDR assessment. Survival probabilities for participants >80 years old at baseline (n=58) were inestimable at T+ time>5 years due to low representation for this age group. Abbreviations: A+ time, estimated years A+ from SILA; CDR, Clinical Dementia Rating; T+ time, estimated years T+ in the meta-temporal ROI from SILA.

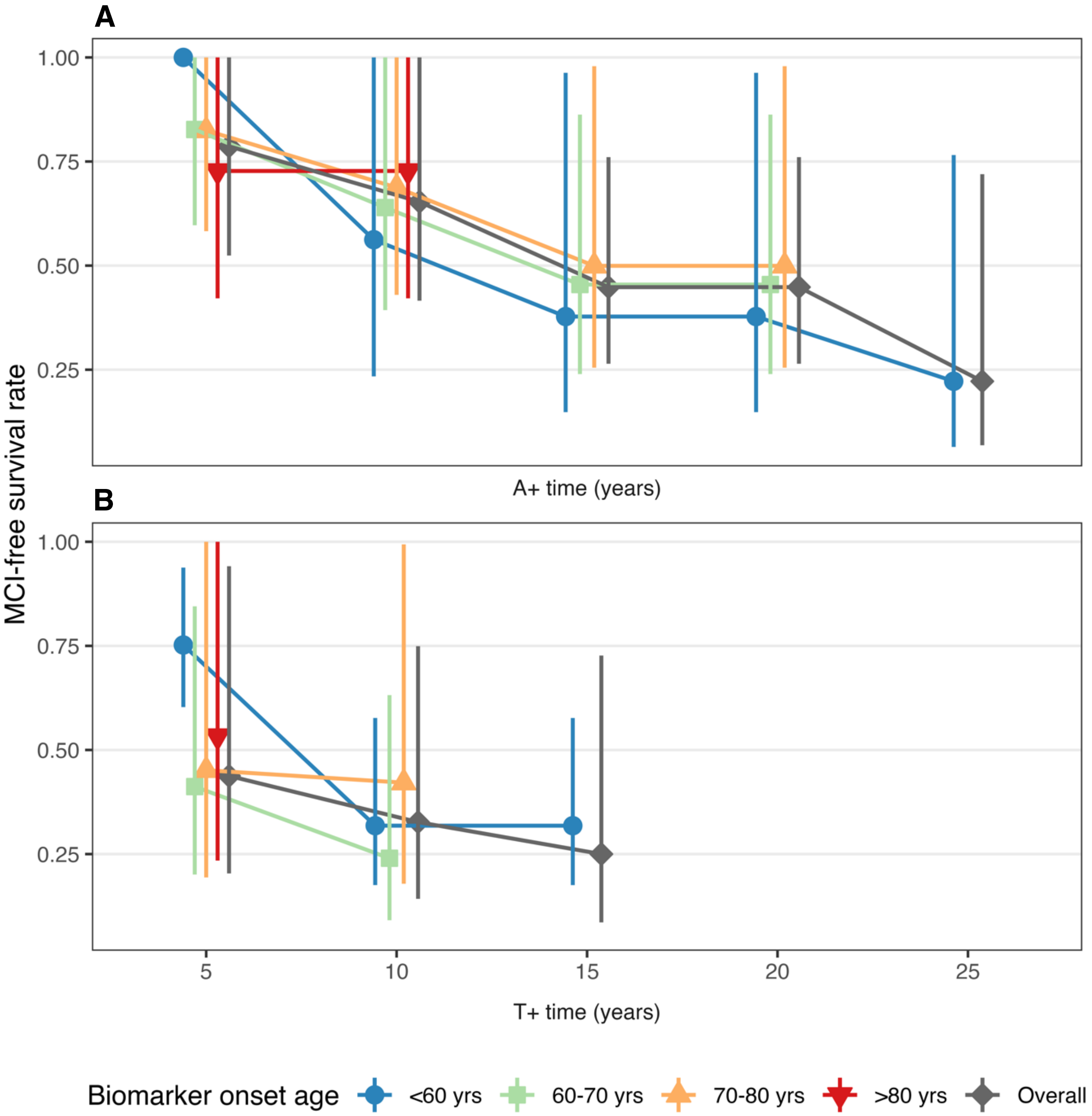


**Supplemental Figure 7. MCI-free survival rates across A+ time and T+ time.** 5- through 25-year MCI-free survival rates after (**A**) A+ onset (n=384 A+) and (**B**) T+ onset (n=190 T+). Individuals were considered to have MCI if they had CDR global scores of 0.5 at two or more consecutive observations. MCI-free probabilities were calculated using accelerated failure time models, and were stratified by decade of EAOA (top) and ETOA (bottom); participants were considered to convert to MCI after two consecutive Clinical Dementia Rating global scores ≥ 0.5. Abbreviations: A+ time, estimated years A+ from SILA; MCI, mild cognitive impairment; T+ time, estimated years T+ in the meta-temporal ROI from SILA.

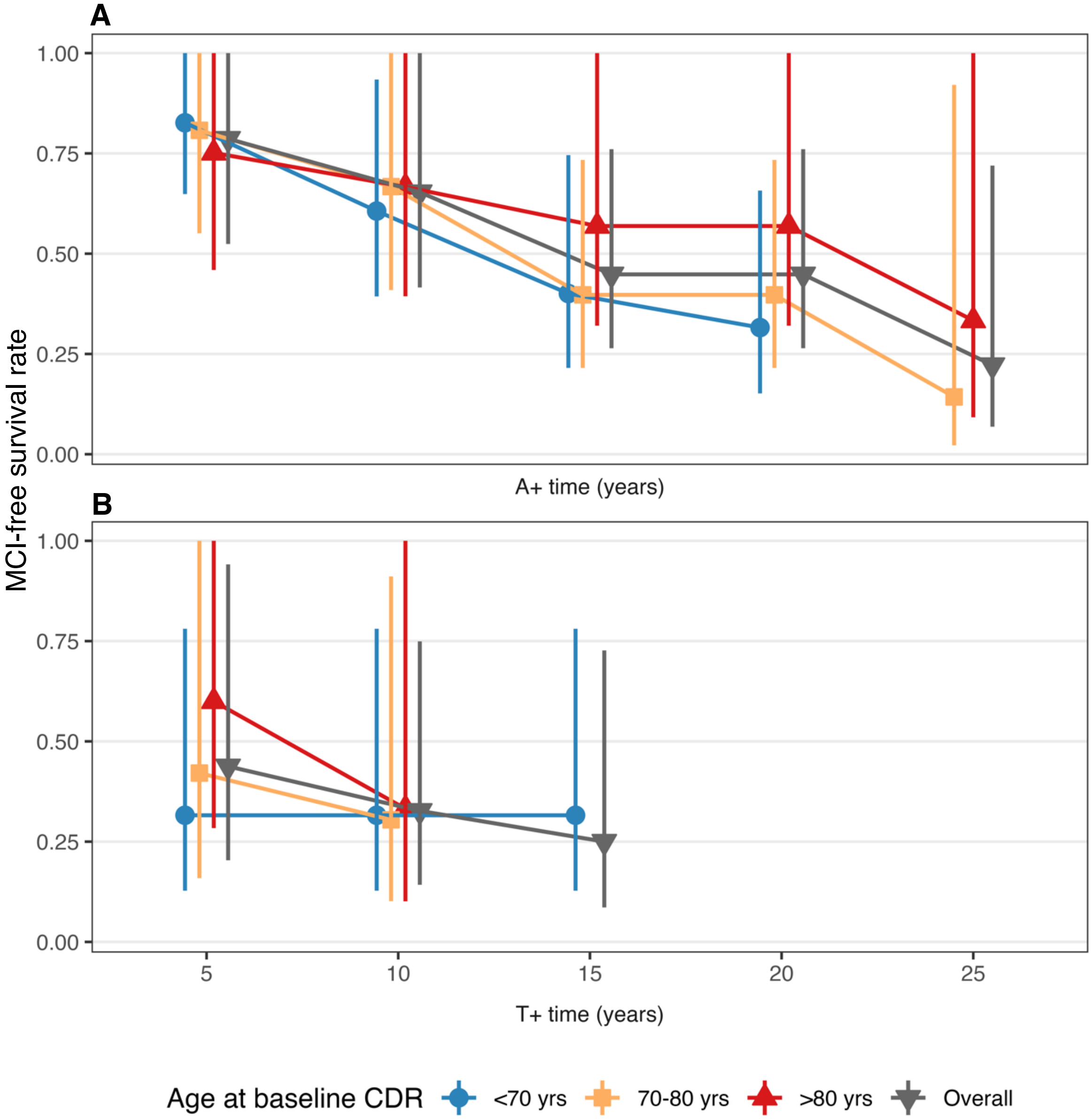


**Supplemental Figure 8. MCI-free survival rates across A+ time and T+ time, stratified by age at baseline CDR.** 5- through 25-year MCI-free survival rates after (**A**) A+ onset (n=384 A+) and (**B**) T+ onset (n=190 T+). MCI-free probabilities were calculated using accelerated failure time models and were stratified by decade of baseline CDR assessment; participants were considered to convert to MCI after two consecutive CDR global scores ≥0.5. Survival probabilities for participants >80 years old at baseline were inestimable at T+ time>10 years. Abbreviations: A+ time, estimated years A+ from SILA; CDR, Clinical Dementia Rating; MCI, mild cognitive impairment; T+ time, estimated years T+ in the meta-temporal ROI from SILA.

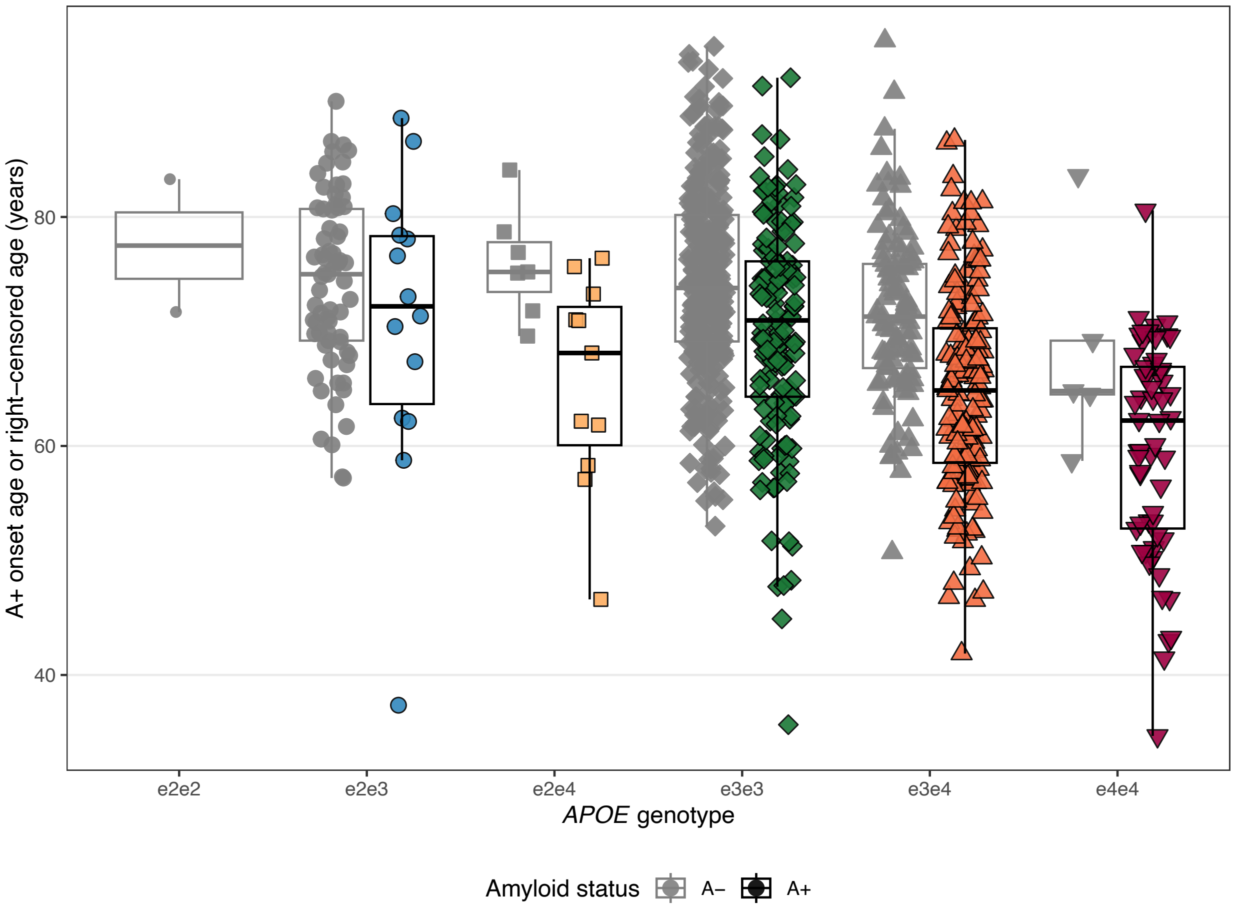


**Supplemental Figure 9. Amyloid onset age as a function of *APOE* genotype.** Estimated age at A+ in the global ROI (Supplementary Figure 1A) illustrated across *APOE* genotypes, shown in the ADNI subset included in the present study to confirm *APOE* effects found in Betthauser *et al.*[3]. Cohort composition in the present study differs due to continued ADNI enrollment and inclusion criteria requiring that participants have tau PET data. Amyloid PET measures also differ due to inclusion of both $^{18}$F-Florbetapir and $^{18}$F-Florbetaben data. Plot color and point shape indicates *APOE* genotype for T+ cases. Age at censor is shown in gray for T-. Abbreviations: A+, amyloid-positive status quantified using $^{18}$F-Florbetapir and $^{18}$F-Florbetaben PET measurements; ADNI, Alzheimer's Disease Neuroimaging Initiative; *APOE*, apolipoprotein E; PET, positron emission tomography.

### Supplemental Tables

**Supplemental Table 1. Aims 3A and 3B demographics compared to Aim 1 full cohort**

| | Cohort | | | p-values | |
|---|---|---|---|---|---|
| **Characteristic** | **Aim 1** N = 905 | **Aim 3A** N = 888 | **Aim 3B** N = 384 | **1 vs. 3A** | **1 vs. 3B** |
| EAOA, mean (SD) years | 66.3 (9.91) | 66.3 (9.91) | 66.4 (9.90) | 0.89* | 0.89* |
| ETOA, mean (SD) years | 72.0 (9.53) | 72.0 (9.63) | 71.5 (9.36) | >0.99* | 0.62* |
| Estimated A at ETOA, mean (SD) CL | 26.4 (42.8) | 25.7 (42.5) | 66.2 (33.7) | 0.73* | <0.001* |
| A+ at or before final A scan, n (%) | 399 (44) | 386 (43) | 384 (100) | 0.79† | <0.001† |
| A+ at or before ETOA, n (%) | 387 (43) | 374 (42) | 367 (96) | 0.78† | <0.001† |
| T+ at or before final T scan, n (%) | 198 (22) | 192 (22) | 175 (46) | 0.90† | <0.001† |
| Age at baseline, mean (SD) years | 71.2 (6.99) | 71.2 (7.03) | 72.7 (6.91) | 0.94* | <0.001* |
| Clinical diagnosis at baseline, n (%) | | | | 0.98† | <0.001† |
| Cognitively unimpaired | 504 (57) | 508 (57) | 165 (43) | | |
| Mild cognitive impairment | 291 (33) | 288 (33) | 145 (38) | | |
| AD dementia | 88 (10.0) | 88 (10.0) | 72 (19) | | |
| Unknown | 22 | 4 | 2 | | |
| Sex, n (%) | | | | 0.79† | 0.60† |
| Male | 441 (49) | 427 (48) | 181 (47) | | |
| Female | 464 (51) | 461 (52) | 203 (53) | | |
| *APOE* genotype, n (%) | | | | >0.99‡ | |
| e2e2 | 2 (0.2) | 2 (0.2) | 0 (0) | | |
| e2e3 | 83 (9.2) | 83 (9.4) | 13 (3.4) | | |
| e2e4 | 18 (2.0) | 18 (2.0) | 11 (2.9) | | |
| e3e3 | 468 (52) | 460 (52) | 145 (38) | | |
| e3e4 | 272 (30) | 261 (30) | 159 (42) | | |
| e4e4 | 62 (6.9) | 58 (6.6) | 53 (14) | | |
| Unknown | 0 | 6 | 3 | | |
| Education level, n (%) | | | | 0.97† | 0.21† |
| High school or less (≤12 years) | 94 (10) | 89 (10) | 42 (11) | | |
| Some college (13-16 years) | 393 (43) | 388 (44) | 185 (48) | | |
| Postgraduate | 418 (46) | 411 (46) | 157 (41) | | |
| Total ANART errors, mean (SD) words | 11.6 (9.74) | 11.5 (9.71) | 12.2 (9.76) | 0.89* | 0.30* |
| Unknown | 21 | 22 | 8 | | |
| Race, n (%) | | | | >0.99‡ | 0.19‡ |
| Asian | 25 (2.8) | 25 (2.8) | 5 (1.3) | | |
| Black or African American | 87 (9.6) | 88 (9.9) | 25 (6.5) | | |
| More than one race | 14 (1.5) | 14 (1.6) | 8 (2.1) | | |
| Native American or Alaskan Native | 2 (0.2) | 2 (0.2) | 0 (0) | | |
| Unknown | 6 (0.7) | 7 (0.8) | 2 (0.5) | | |
| White | 771 (85) | 752 (85) | 344 (90) | | |
| Ethnicity, n (%) | | | | 0.95‡ | 0.62‡ |
| Hispanic or Latino | 57 (6.3) | 58 (6.5) | 20 (5.2) | | |
| Not Hispanic or Latino | 845 (93) | 827 (93) | 362 (94) | | |
| Unknown | 3 (0.3) | 3 (0.3) | 2 (0.5) | | |

Participants were included in Aim 3A to examine heterogeneity in cognitive performance with respect to amyloid and tau timelines. Aim 3A cohort included the subset of Aim 1 participants who had CDR data available. Participants were included in Aim 3B to calculate the 5- through 25-year dementia-free survival rates among A+ individuals and compare against survival rates among T+ individuals (n=190; demographics characterized in main text Table 1, cohort header Aim 2). EAOA and ETOA are calculated among A+ and T+ participants, respectively. Abbreviations: A+, amyloid positivity assessed using $^{18}$F-Florbetapir or $^{18}$F-Florbetaben PET neuroimaging; AD, Alzheimer's disease; ANART, American National Adult Reading Test; *APOE*, apolipoprotein E; CL, Centiloids; EAOA, estimated amyloid onset age; ETOA, estimated tau onset age; SD, standard deviation; T+, tau positivity assessed using $^{18}$F-Flortaucipir PET neuroimaging.

*One-way analysis of means (not assuming equal variances)

†Pearson's Chi-squared test

‡Fisher's exact test

**Supplemental Table 2. Forward selection of biological variables predicting T+ risk in Cox proportional hazards models**

| Characteristic | N=885 | Event N | Model 1.1<br>Sex<br>HR (95% CI)[1,2] | Model 1.2<br>+ APOE<br>HR (95% CI)[1,2] | Model 1.3<br>+ A at ETOA<br>HR (95% CI)[1,2] | Model 1.4<br>+ Age at baseline FTP<br>HR (95% CI)[1,2] | Model 1.5<br>+ Sex x APOE<br>HR (95% CI)[1,2] | Model 1.6<br>+ Sex x A at ETOA<br>HR (95% CI)[1,2] | Model 1.7<br>+ Sex x APOE<br>+ Sex x A at ETOA<br>HR (95% CI)[1,2] |
|---|---|---|---|---|---|---|---|---|---|
| Sex (Male reference) | 432 | 88 | — | — | — | — | — | — | — |
| Female | 453 | 104 | 1.503<br>(1.129-2.000)** | 1.651<br>(1.235-2.206)*** | 1.813<br>(1.349-2.436)*** | 1.288<br>(0.956-1.735) | 1.104<br>(0.664-1.835) | 0.829<br>(0.501-1.372) | 0.806<br>(0.438-1.482) |
| *APOE* genotype (ε3ε3 reference) | 468 | 60 | | — | — | — | — | — | — |
| ε2ε3 | 83 | 6 | | 0.628<br>(0.271-1.454) | 0.871<br>(0.373-2.032) | 0.843<br>(0.361-1.967) | 1.289<br>(0.448-3.706) | 0.825<br>(0.354-1.927) | 1.114<br>(0.385-3.223) |
| ε3ε4 | 272 | 94 | | 3.733<br>(2.691-5.180)*** | 2.623<br>(1.871-3.679)*** | 1.783<br>(1.265-2.514)*** | 1.411<br>(0.865-2.301) | 1.822<br>(1.288-2.576)*** | 1.544<br>(0.935-2.549) |
| ε4ε4 | 62 | 32 | | 7.577<br>(4.903-11.71)*** | 3.888<br>(2.451-6.168)*** | 2.713<br>(1.700-4.331)*** | 2.93<br>(1.549-5.541)*** | 2.76<br>(1.722-4.421)*** | 3.293<br>(1.713-6.331)*** |
| Estimated A at ETOA (10 CL) | 885 | 192 | | | 1.133<br>(1.097-1.170)*** | 1.206<br>(1.166-1.249)*** | 1.208<br>(1.167-1.251)*** | 1.164<br>(1.110-1.221)*** | 1.169<br>(1.113-1.228)*** |
| Age at baseline FTP (10 years) | 885 | 192 | | | | 0.122<br>(0.086-0.173)*** | 0.121<br>(0.085-0.172)*** | 0.121<br>(0.085-0.172)*** | 0.119<br>(0.084-0.170)*** |
| Sex x *APOE* (Female ε3ε3 reference) | 236 | 30 | | | | | | | |
| Female x ε2ε3 | 41 | 2 | | | | | 0.395<br>(0.067-2.339) | | 0.492<br>(0.082-2.954) |
| Female x ε3ε4 | 145 | 56 | | | | | 1.531<br>(0.792-2.959) | | 1.338<br>(0.681-2.629) |
| Female x ε4ε4 | 31 | 16 | | | | | 0.871<br>(0.362-2.095) | | 0.724<br>(0.295-1.776) |
| Female x CL at ETOA | 453 | 104 | | | | | | 1.068<br>(1.005-1.134)* | 1.061<br>(0.995-1.132) |
| Log-likelihood | | | -1,137 | -1,080*** | -1,051*** | -966*** | -964 | -964 | -962 |
| AIC | | | 2,276 | 2,168 | 2,112 | 1,944 | 1,946 | 1,942 | 1,944 |
| BIC | | | 2,279 | 2,181 | 2,128 | 1,964 | 1,975 | 1,964 | 1,977 |

p<0.1; *p<0.05; **p<0.01; ***p<0.001. Cox proportional hazards models were applied to estimate the hazard ratio of progressing to T+; forward selection was used to identify biological factors that significantly contributed to outcome variance (Log-ratio *P*<.05). Model comparisons for likelihood ratio tests: 2 vs 1, 3 vs 2, 4 vs 3, 5 vs 4, 6 vs 5. Model 1.4 (outlined in red) explained more variance in the ETOA event compared to Model 1.3 (log-likelihood *P*<2.2e-16). This resulted in a base model including the predictors Sex + *APOE* + CL at ETOA + baseline age. We additionally tested Models 1.5, 1.6, and 1.7 per prespecified exploratory analyses. Abbreviations: AIC, Akaike Information Criterion; BIC, Bayesian Information Criterion; CI, confidence interval; CL at ETOA, estimated amyloid burden at ETOA; ETOA, estimated tau onset age; FTP, $^{18}$F-Flortaucipir; HR, hazard ratio.

**Supplemental Table 3. Literacy exposure and education predictors of T+ risk in Cox proportional hazards models**

| | | | Model 1.4 | Model 1.8 | Model 1.9 | Model 1.10 |
|---|---|---|---|---|---|---|
| | | | Base Model | ANART errors | Education | ANART + education |
| **Characteristic** | **N=864** | **Event N** | **HR (95% CI)** | **HR (95% CI)** | **HR (95% CI)** | **HR (95% CI)** |
| Sex (male reference) | 419 | 84 | — | — | — | — |
| Female | 445 | 102 | 1.331 (0.982-1.804) | 1.374 (1.013-1.865)* | 1.242 (0.916-1.685) | 1.302 (0.957-1.771) |
| *APOE* genotype (ε3ε3 reference) | 453 | 57 | — | — | — | — |
| ε2ε3 | 82 | 6 | 0.87 (0.372-2.035) | 0.818 (0.349-1.915) | 0.842 (0.359-1.974) | 0.823 (0.351-1.929) |
| ε3ε4 | 267 | 91 | 1.785 (1.257-2.536)** | 1.869 (1.317-2.653)*** | 1.835 (1.293-2.604)*** | 1.888 (1.332-2.676)*** |
| ε4ε4 | 62 | 32 | 2.771 (1.728-4.444)*** | 2.817 (1.755-4.522)*** | 2.947 (1.836-4.728)*** | 2.967 (1.848-4.766)*** |
| Estimated A at ETOA (10 CL) | 864 | 186 | 1.207 (1.166-1.250)*** | 1.201 (1.159-1.243)*** | 1.198 (1.157-1.241)*** | 1.195 (1.154-1.238)*** |
| Age at baseline FTP (10 years) | 864 | 186 | 0.118 (0.083-0.169)*** | 0.114 (0.079-0.163)*** | 0.118 (0.083-0.169)*** | 0.115 (0.080-0.164)*** |
| ANART errors | 864 | 186 | | 1.022 (1.009-1.036)*** | | 1.019 (1.004-1.034)* |
| Education (college reference) | 374 | 99 | | | — | — |
| High school or less | 90 | 24 | | | 0.986 (0.629-1.547) | 0.826 (0.516-1.322) |
| Postgraduate | 400 | 63 | | | 0.623 (0.450-0.863)** | 0.685 (0.490-0.958)* |
| Log-likelihood | | | -928 | -923** | -924* | -921** |
| AIC | | | 1,868 | 1,860 | 1,863 | 1,859 |
| BIC | | | 1,888 | 1,883 | 1,889 | 1,888 |

.p<0.1; *p<0.05; **p<0.01; ***p<0.001. Educational experiences coefficients are calculated using the base model (Model 1.4) plus educational experiences variables as listed in the column headers, including the complete cases in the dataset (i.e., those with ANART and education data available). All models' log-likelihood values were compared to Model 1.4. Education attainment levels are defined as such: high school or less (≤12 years), college (13-16 years), postgraduate (>16 years). Abbreviations: ANART, American National Adult Reading Test. AIC, Akaike Information Criterion; BIC, Bayesian Information Criterion; CI, confidence interval; CL at ETOA, estimated amyloid burden at ETOA; ETOA, estimated tau onset age; FTP, $^{18}$F-Flortaucipir; HR, hazard ratio.

### Supplemental Table 4. Biological predictors of time from T+ onset to dementia in the T+ subset using accelerated failure time models

| | | | Model 2.1 | Model 2.2 | Model 2.3 | Model 2.4 | Model 2.5 | Model 2.6 | Model 2.7 | Model 2.8 |
|---|---|---|---|---|---|---|---|---|---|---|
| | | | A at ETOA | + ETOA | + APOE | + Sex | + Age at baseline FTP | + Sex x APOE | + Sex x A at ETOA | + Sex x APOE + Sex x A at ETOA |
| **Characteristic** | **N= 178** | **Event N** | **Beta (95% CI)** | **Beta (95% CI)** | **Beta (95% CI)** | **Beta (95% CI)** | **Beta (95% CI)** | **Beta (95% CI)** | **Beta (95% CI)** | **Beta (95% CI)** |
| Estimated A at ETOA (10 CL) | 178 | 114 | -0.071 (-0.113--0.030)*** | -0.046 (-0.079--0.012)** | -0.043 (-0.078--0.009)* | -0.045 (-0.079--0.011)** | -0.044 (-0.078--0.009)* | -0.041 (-0.075--0.007)* | -0.045 (-0.081--0.008)* | -0.023 (-0.068-0.021) |
| ETOA (10 years) | 178 | 114 | | -0.286 (-0.421--0.152)*** | -0.294 (-0.435--0.152)*** | -0.284 (-0.417--0.150)*** | -0.351 (-0.743-0.042) | -0.293 (-0.436--0.150)*** | -0.286 (-0.421--0.152)*** | -0.281 (-0.424--0.138)*** |
| *APOE* genotype (ε3ε3 reference) | 59 | 41 | | | — | | | — | | — |
| ε3ε4 | 90 | 48 | | | 0.04 (-0.236-0.316) | | | 0.138 (-0.235-0.510) | | 0.142 (-0.226-0.511) |
| ε4ε4 | 29 | 25 | | | -0.099 (-0.427-0.229) | | | 0.087 (-0.336-0.510) | | 0.102 (-0.318-0.522) |
| Sex (Male reference) | 82 | 55 | | | | — | | — | | — |
| Female | 96 | 59 | | | | 0.054 (-0.176-0.283) | | 0.209 (-0.182-0.600) | | 0.508 (-0.138-1.154) |
| Age at baseline FTP (10 years) | 178 | 114 | | | | | 0.085 (-0.404-0.575) | | | |
| Sex x *APOE* (Female ε3ε3 reference) | 178 | 114 | | | | | | — | | — |
| Female x ε3ε4 | 53 | 28 | | | | | | -0.189 (-0.716-0.338) | | -0.226 (-0.753-0.301) |
| Female x ε4ε4 | 14 | 13 | | | | | | -0.415 (-1.038-0.209) | | -0.41 (-1.026-0.207) |
| Female x A at ETOA (10 CL) | 96 | 59 | | | | | | | -0.001 (-0.029-0.027) | -0.038 (-0.104-0.028) |
| Log-likelihood | | | -192 | -185*** | -184 | -185 | -185 | -184 | -185 | -183 |
| AIC | | | 390 | 378 | 381 | 379 | 380 | 385 | 380 | 386 |
| BIC | | | 400 | 390 | 400 | 395 | 395 | 414 | 396 | 418 |

*p<0.05; **p<0.01; ***p<0.001. Model comparisons for likelihood ratio tests: 2 vs 1, 3 vs 2, 4 vs 3, 5 vs 4, 6 vs 5, 7 vs 6. Accelerated failure time model coefficients reflect the differences in years from T+ to dementia between each variable stratum and its reference group. Compared to ETOA at 60 years old, for instance, participants with ETOA at 70 years old converted to dementia 29% faster after T+ onset. Model 2.2 (outlined in red) explained more variance in the time from ETOA to dementia onset age, compared to Model 2.1 (log-likelihood *p*=1.3e-4); Models 2.3 through 2.5 did not explain additional variance. This resulted in a base model including the predictors CL at ETOA + ETOA. Effects of A at ETOA and female sex x A at ETOA are calculated for 10 CL amyloid increases. Abbreviations: AIC, Akaike Information Criterion; BIC, Bayesian Information Criterion; CI, confidence interval; CL at ETOA, estimated amyloid burden at ETOA; ETOA, estimated tau onset age; FTP, $^{18}$F-Flortaucipir.

**Supplemental Table 5. Literacy exposure and education effects on time from T+ onset to dementia in the T+ subset using accelerated failure time models**

| | | | Model 2.2 | Model 2.9 | Model 2.10 | Model 2.11 |
|---|---|---|---|---|---|---|
| | | | Base model | ANART | Education | ANART + education |
| **Characteristic** | **N** | **Event N** | **Beta (95% CI)** | **Beta (95% CI)** | **Beta (95% CI)** | **Beta (95% CI)** |
| Estimated A at ETOA (10 CL) | 172 | 112 | -0.045 (-0.079--0.011)** | -0.044 (-0.078--0.010)* | -0.04 (-0.074--0.006)* | -0.041 (-0.075--0.007)* |
| ETOA (10 years) | 172 | 112 | -0.28 (-0.415--0.145)*** | -0.28 (-0.415--0.144)*** | -0.296 (-0.431--0.161)*** | -0.29 (-0.427--0.153)*** |
| ANART errors | 172 | 112 | | -0.011 (-0.023-0.000) | | -0.01 (-0.022-0.002) |
| Education (college reference) | 94 | 64 | | | — | — |
| High school or less | 19 | 16 | | | 0.037 (-0.301-0.375) | 0.079 (-0.262-0.421) |
| Postgraduate | 59 | 32 | | | 0.186 (-0.079-0.451) | 0.138 (-0.131-0.408) |
| Log-likelihood | | | -182 | -180 | -181 | -180 |
| AIC | | | 372 | 371 | 374 | 374 |
| BIC | | | 385 | 386 | 393 | 396 |

*p<0.05; **p<0.01; ***p<0.001. Accelerated failure time model coefficients reflect the differences in years from T+ to dementia between each variable stratum and its reference group. Compared to ETOA at 60 years old, for instance, participants with ETOA at 70 years old converted to dementia 29% faster after T+ onset. Educational experiences coefficients are calculated using the base model (Model 2.2) plus educational experiences variables as listed in the column headers, including the complete cases in the dataset (i.e., those with ANART and education data available). Education attainment levels are defined as such: high school or less (≤12 years), college (13-16 years), postgraduate (>16 years). Abbreviations: ANART, American National Adult Reading Test. AIC, Akaike Information Criterion; BIC, Bayesian Information Criterion; CI, confidence interval; CL at ETOA, estimated amyloid burden at ETOA; ETOA, estimated tau onset age; FTP, $^{18}$F-Flortaucipir

### References


[1] Landau SM, Fero A, Baker SL, Koeppe R, Mintun M, Chen K, et al. Measurement of longitudinal β-amyloid change with 18F-florbetapir PET and standardized uptake value ratios. J Nucl Med Off Publ Soc Nucl Med 2015;56:567–74. https://doi.org/10.2967/jnumed.114.148981.

[2] for the Alzheimer’s Disease Neuroimaging Initiative, Royse SK, Minhas DS, Lopresti BJ, Murphy A, Ward T, et al. Validation of amyloid PET positivity thresholds in centiloids: a multisite PET study approach. Alzheimers Res Ther 2021;13:99. https://doi.org/10.1186/s13195-021-00836-1.

[3] Betthauser TJ, Bilgel M, Koscik RL, Jedynak BM, An Y, Kellett KA, et al. Multi-method investigation of factors influencing amyloid onset and impairment in three cohorts. Brain 2022;145:4065–79. https://doi.org/10.1093/brain/awac213.
